# A Critical Analysis of UK Media Characterisations of Long Covid in Children and Young People

**DOI:** 10.1101/2024.04.13.24305152

**Authors:** Chloe Connor, Michael Kranert, Sara Mckelvie, Donna Clutterbuck, Sammie Mcfarland, Nisreen A Alwan

## Abstract

Long Covid is the continuation or development of symptoms related to a SARSCoV2 infection. Those with Long Covid may face epistemic injustice, where they are unjustifiably viewed as unreliable evaluators of their own illness experiences. Media articles both reflect and influence perception and subsequently how people regard children and young people (CYP) with Long Covid, and may contribute to epistemic injustice.

We aimed to explore how the UK media characterises Long Covid in CYP through examining three key actor groups: parents, healthcare professionals, and CYP with Long Covid, through the lens of epistemic injustice. A systematic search strategy resulted in the inclusion of 103 UK media articles. We used an adapted corpus-assisted Critical Discourse Analysis in tandem with thematic analysis. Specifically, we utilised search terms to locate concordances of key actor groups.

In the corpus, parents highlighted minimisation of Long Covid, barriers to care, and experiences of personal attacks. Mothers were presented as also having Long Covid. Fathers were not mentioned once. Healthcare professionals emphasised the rarity of Long Covid in CYP, avoided pathologizing Long Covid, and overemphasised psychological components. CYP rarely were consulted in media articles but were presented as formerly very able. Manifestations of Long Covid in CYP were validated or invalidated in relation to adults.

Media characterisations contributed to epistemic injustice. The disempowering portrayal of parents promote stigma and barriers to care. Healthcare professionals’ narratives often contributed to negative healthcare experiences and enacted testimonial injustice, where CYP and parent’s credibility was diminished due to unfair identity prejudice, in their invalidation of Long Covid. Media characterisations reveal and maintain a lack of societal framework for understanding Long Covid in CYP. The findings of this study illustrate the discursive practices employed by journalists that contribute to experiences of epistemic injustice. Based on our findings, we propose recommendations for journalists.

## Introduction

Long Covid in children and young people (CYP) occurs in those with a history of confirmed or probable SARS-CoV-2 infection, with symptoms lasting at least 2 months initially occurring within 3 months of an acute covid-19 infection.^1^ Potential symptoms range widely and include cognitive difficulties, cough, dizziness, dyspnoea, joint pain, light sensitivity, loss of appetite, myalgia, palpitations, and sore eyes or throat, and can newly onset or persist from the initial infection. The World Health Organisation (WHO) definition of Long Covid in CYP was developed in February 2023 to align understanding of the condition and acknowledge that CYP have potentially different Long Covid presentations from adults.^1^

Long Covid is the first illness to be socially constructed through afflicted individuals connecting online.^2^ While the developing understanding of Long Covid has more patient input than seen in other diseases, people living with Long Covid nevertheless experience barriers to recognition of their experience and perspectives.^3^ In addition to the requirement of proof of infection, there are formidable barriers for those with Long Covid, especially CYP, to accessing adequate care.^4^ Long Covid services often require a general practitioner (GP) referral,^5,6^ and many clinics continue to have a wait time of over 15 weeks.^7^ In addition to the logistic barriers to care, people with Long Covid face discrimination and stigma which hinders engagement with health services and can result in healthcare professionals (HCPs) minimising the experience of people with Long Covid.^4–6,8^

Epistemic injustice occurs when people are unjustifiably discredited, as unreliable evaluators of their own illness experiences.^9^ There are two forms of epistemic injustice: testimonial and hermeneutical.^10^ Testimonial injustice occurs when someone’s credibility is diminished because of unfair identity prejudice.^10^ This has been seen in Long Covid, where lived experiences are dismissed due to those living or describing them being negatively stereotyped.^11^ These negative stereotypes can be formed by aspects of a person’s identity that unfairly diminish their perceived credibility, such as their race, gender, social class, or age.

The other form of epistemic injustice is hermeneutic injustice, where a person is not able to articulate their experience because of a gap in collective interpretive resources.^12^ The hermeneutic injustice in Long Covid stems from a societal lack of a framework for understanding and conceptualising the condition. There is still limited understanding of Long Covid partially due to its relatively recent emergence, and this hinders recognition of the condition. The predominance of the biomedical illness model for conceptualising disease in countries such as the UK privileges diseases diagnosable by an “objective” test over diseases that are predominately diagnosed via symptom presentation.^11^ There is still no biomarker that can offer sensitive and specific diagnosis of Long Covid. As a result, Long Covid suffers low disease prestige and those afflicted are disadvantaged by this.^11^

Systemic power and social structures influence the characterisation of Long Covid.^10^ The media play a large role in the knowledge construction of certain chronic diseases and the epistemic (in)justice in representing various actors involved.^10^ Media articles both reflect and influence perception of the condition and subsequently how people regard and behave towards CYP with Long Covid. Key actors such as HCPs, parents of children with Long Covid, and affected CYP are frequently represented in media articles reporting on Long Covid in CYP. In the articles, the actors share their knowledge and are also discussed by others. There is currently a lack of research analysing media coverage of Long Covid. We aimed to examine how the UK media characterise Long Covid in CYP using a Critical Discourse Analysis approach.

## Methods

### Data sources and systematic search strategy

This study analyses media articles about Long Covid in CYP published in national newspapers in the UK between January 1st, 2020 and June 7th, 2023. Articles were collected through the search engine LexisNexis using search terms related to Long Covid and CYP. After duplicates were removed and all articles were skimmed for relevance according to the inclusion criteria (**Table 1**), 103 articles were included for analysis. The adapted PRISMA diagram is presented in **Fig 1**. For the full systematic search strategy, a list of included/excluded publishers, descriptive data including a demographic breakdown of the included articles (style and political leaning of publisher, date published), refer to **SI 1 and 2 tables, SI 3-5 figures.** For the characterisation of each publisher, refer to **SI 6 table**. Articles were labelled as duplicates if they were published within 48 hours with the same author(s), with an identical or nearly identical text body. Articles with repeated text but significantly different lengths (as assessed by the primary researcher) were not considered duplicates.

**Figure 1:**
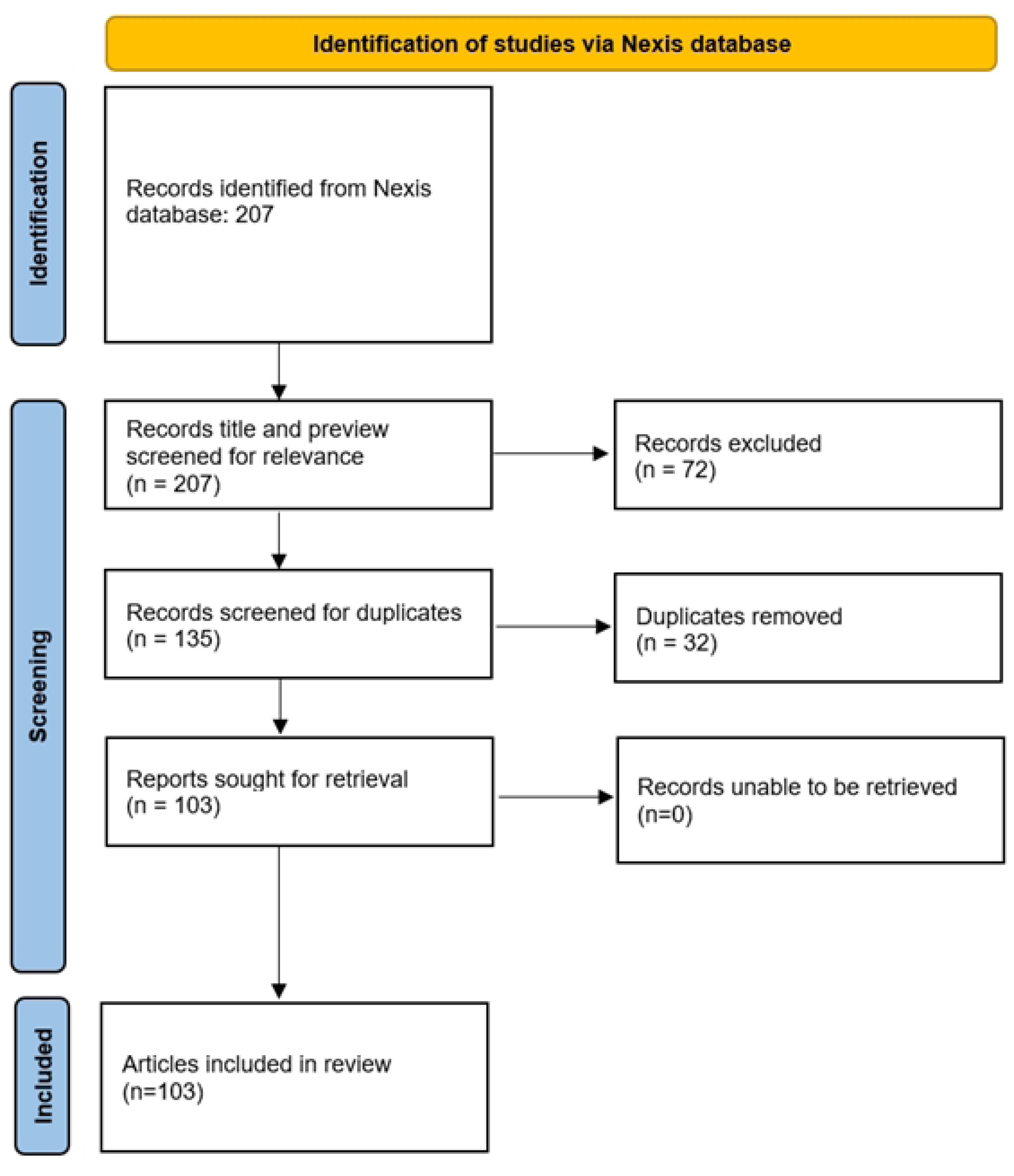
Adapted PRISMA diagram of media articles identified for inclusion.

**Figure 2.**
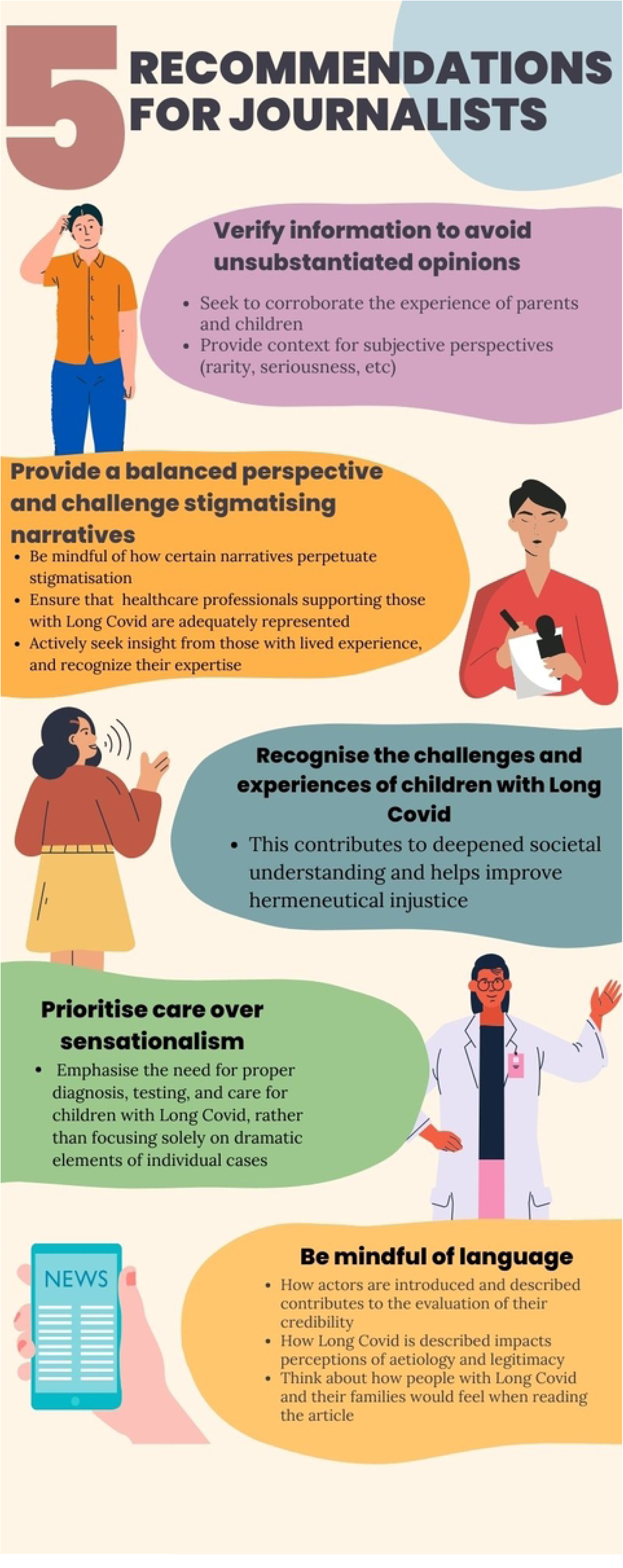
Recommendations for journalists to counter epistemic injustice in reporting Long Covid and similar conditions. Graphic created using Canva software.

**Table 1.**
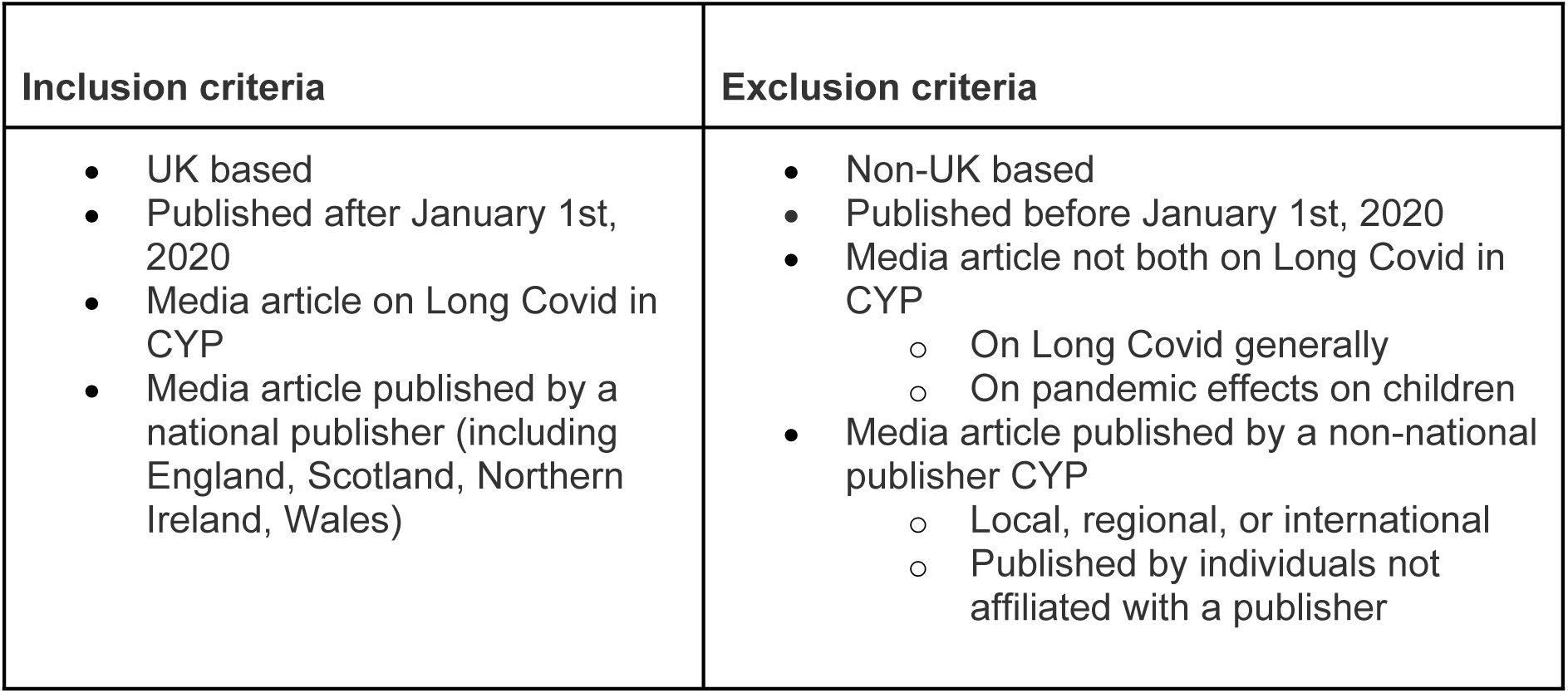
Inclusion and exclusion criteria for media articles.

| Inclusion criteria | Exclusion criteria |
| --- | --- |
| • UK based<br>• Published after January 1st, 2020<br>• Media article on Long Covid in CYP<br>• Media article published by a national publisher (including England, Scotland, Northern Ireland, Wales) | • Non-UK based<br>• Published before January 1st, 2020<br>• Media article not both on Long Covid in CYP<br>○ On Long Covid generally<br>○ On pandemic effects on children<br>• Media article published by a non-national publisher CYP<br>○ Local, regional, or international<br>○ Published by individuals not affiliated with a publisher |

**Table 2.**
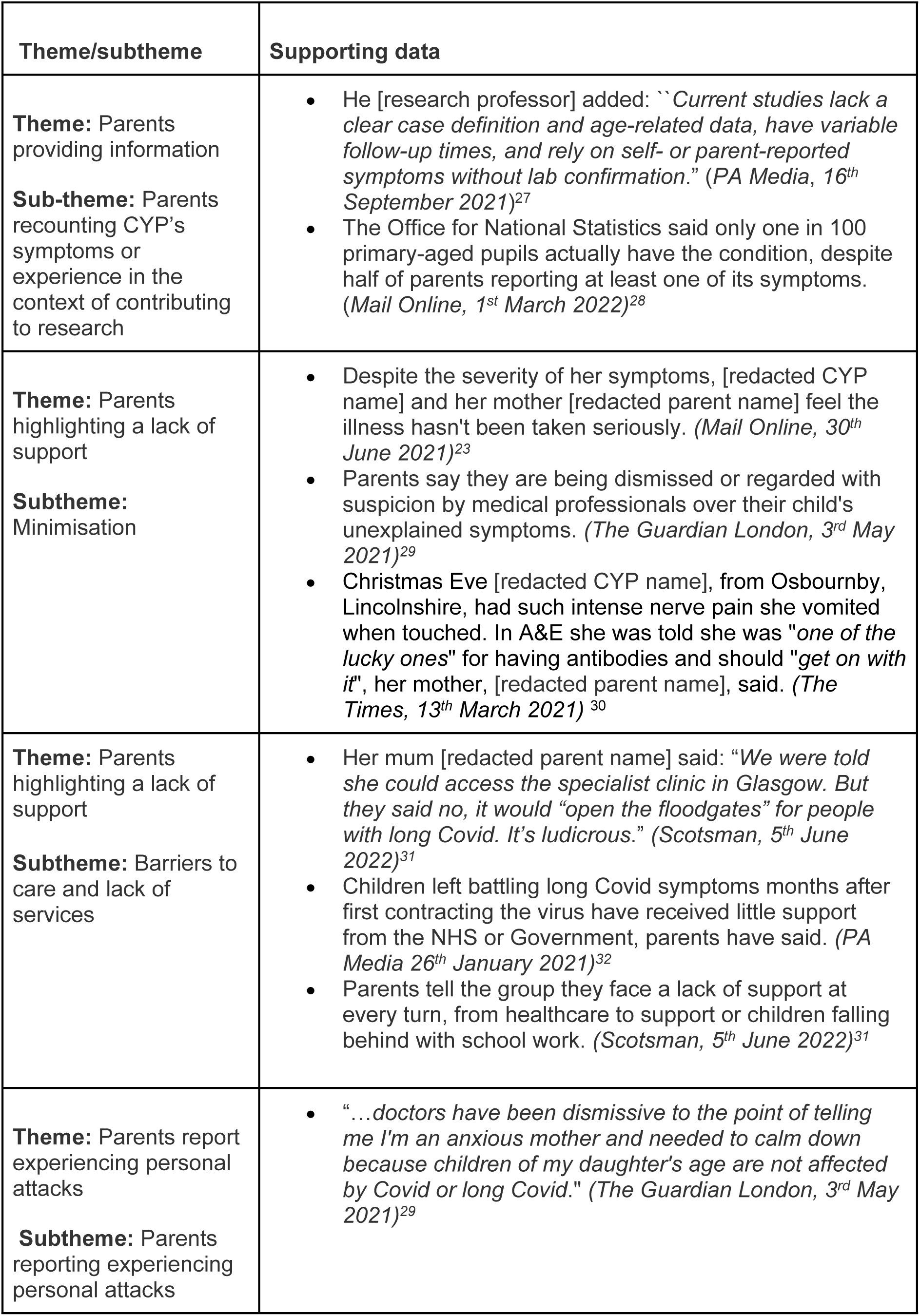
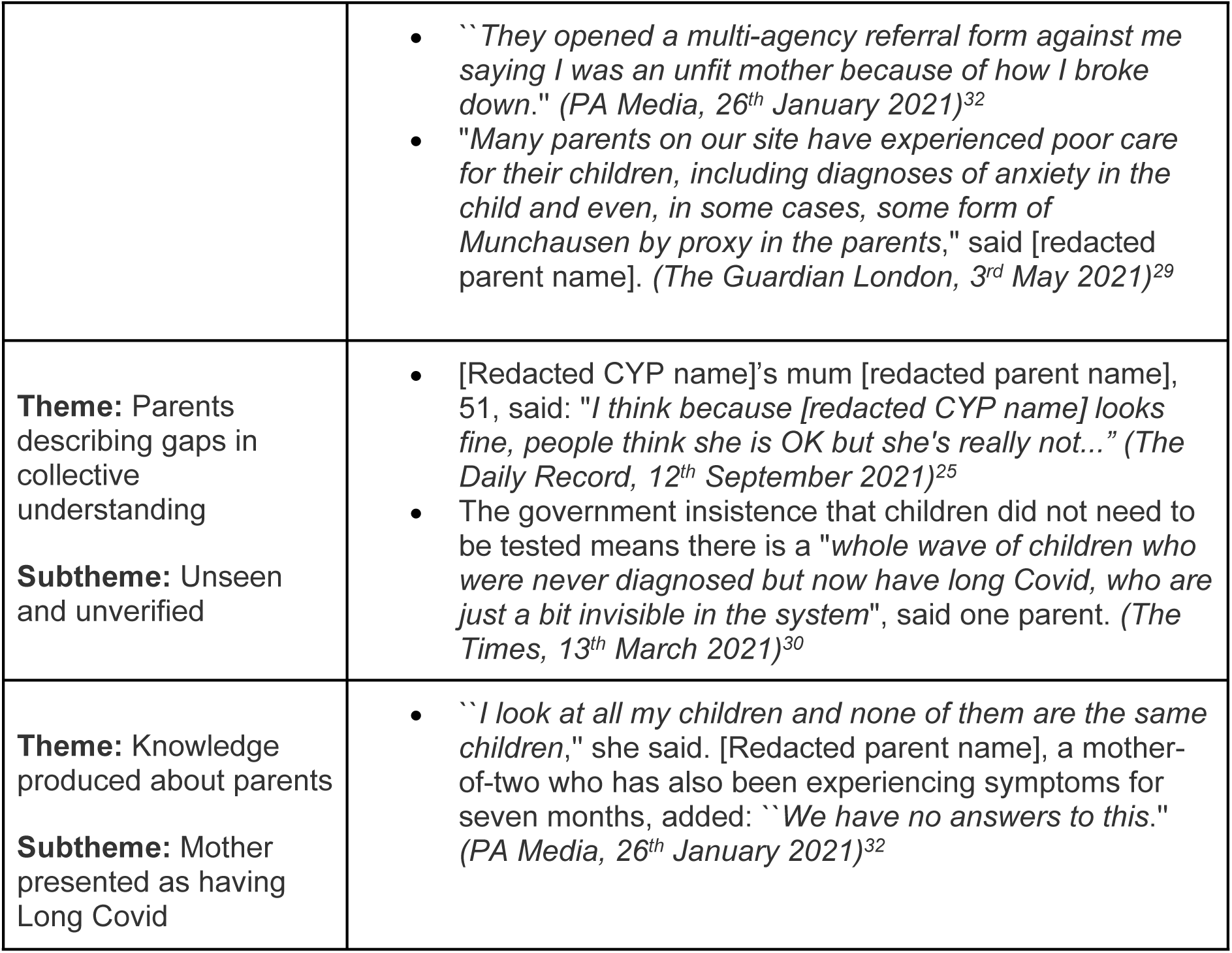
Themes for parents.

**Table 3:** Themes for HCPs.

| Theme/Subtheme | Supporting data |
| --- | --- |
| **Theme:** rarity discourse from HCPs<br>**Subtheme:** Long Covid is rare | • Dr [redacted HCP name], consultant paediatrician at the UK Health Security Agency and study chief investigator, said: *'It is reassuring that the vast majority of primary and secondary school aged children surveyed since March 2020 have not experienced long Covid symptoms. 'These new data should be reassuring for parents, clinicians and policy-makers. (Mail Online, 1<sup>st</sup> March, 2022)<sup>28</sup>*<br>• Dr [redacted HCP name], based at Mater, UCD and Rotunda hospitals, warned: *"Long Covid seems to be rare in children, but it doesn't matter. If it's your child there needs to be a service for even one child with Long Covid. It's a failure once again." (The Sun, 15<sup>th</sup> August, 2022)<sup>34</sup>* |
| **Theme:** HCP perceptions of disease validity<br>**Subtheme:** Long Covid is validated | • [Redacted HCP name] warned: *'Kids get less Covid symptoms, they are less likely to die, they are less likely to end up hospital patients. But they do get side effects.'* (The Scottish Daily Mail, 22<sup>nd</sup> July 2021)<sup>35</sup><br>• GP and author Dr [redacted HCP name] said that while the virus was mild in young children, they were getting long Covid, which was a *"real concern"*. (The Daily Mirror, 14<sup>th</sup> March 2022)<sup>38</sup> |
| **Theme:** HCP perceptions of disease validity<br>**Subtheme:** Pathologisation avoidance | • Dr [redacted HCP name] of the MCRI and University of Fribourg said symptoms of long Covid were difficult to distinguish from those attributable to the indirect effects of the pandemic, such as school closures, not seeing friends or being unable to do sports and hobbies. (PA Media, 16<sup>th</sup> September 2021)<sup>27</sup><br>• Dr [redacted HCP name] said long Covid exhibits the same pattern as other post-viral illnesses, which children are as susceptible to, as adults. Most people will experience some level of post-viral fatigue at some point in their lives. (The Daily Mirror, 1<sup>st</sup> November 2020)<sup>39</sup><br>• *"I'm talking to paediatricians who are already getting referrals - the numbers aren't huge ... I don't think there's a huge cause for concern,"* she said, adding that what parents are most frustrated by is that nobody knows much about it because it's a new condition. *"The good news is that the majority of young people who get chronic fatigue tend to get better with appropriate support."* (The Guardian, 2<sup>nd</sup> March 2021)<sup>40</sup> |
| **Theme:** HCP perceptions of disease validity<br>**Subtheme:** Long Covid as an already established psychological disorder | • Other doctors had determined that her condition was psychological. (*The Guardian*, 10<sup>th</sup> August 2021)<sup>41</sup><br>• The study, which has been running since March 2020, involved 134 schools and inputs from the parents of 4,870 pupils. Dr [redacted HCP name], of King's College London, said: *"There was no significant difference in the numbers presenting with a 'probable mental disorder' between both groups, whether test positive or negative..."* (*The Daily Telegraph*, 1<sup>st</sup> March 2022)<sup>36</sup><br>• Kids with long Covid are treated terribly. The failings of doctors on this is huge. Most still put it down to anxiety. (*The Scotsman*, 5<sup>th</sup> June 2022)<sup>31</sup> |
| **Theme:** Difficulties with diagnosis<br>**Subtheme:** difficulties with diagnosis | • Dr [redacted HCP name], a GP and Glasgow Tory MSP, has raised concerns that long Covid in children is that it can be particularly difficult to diagnose. He said: *'The problem with kids is that, unless it's blindingly obvious, it's difficult getting information out of them.'* (*The Scottish Daily Mail*, 22<sup>nd</sup> July 2021)<sup>35</sup><br>• Dr [redacted HCP name], a champion for Long Covid Kids Scotland, told Scotland on Sunday: *"Long Covid in kids seems to be hidden but the data shows it's a big problem. My concern is what happens if we don't get confirmation of infection. This is so important. It will have an impact and will increase inequalities. Those who can pay will access tests."* (*The Scotsman*, 5<sup>th</sup> June 2022)<sup>31</sup> |
| **Theme:** HCPs uninformed | • *"From the start we have been doing this blind. Doctors have no strategy for how to help [redacted CYP name]..."* (*The Scotsman*, 5<sup>th</sup> June 2022)<sup>31</sup><br>• *"Our children aren't being recognised as Long Covid sufferers because doctors aren't joining the dots between a wide range of symptoms."* (*The Sun*, 7<sup>th</sup> February 2021)<sup>26</sup><br>• Her mother, [redacted parent name], said clinicians have been supportive, but they have *"openly admitted they don't know a lot about long Covid"*. (*Independent Print*, 16<sup>th</sup> June 2021)<sup>42</sup><br>• *"GPs deal in certainties but there are no certainties here, just a litany of new symptoms. We've been bounced endlessly between child and mental health services and the GP, but still no one has any idea how to help my son."* (*The Guardian*, 3<sup>rd</sup> May 2021)<sup>43</sup> |

**Table 4:** Themes for CYP.

| Theme/Subtheme | Supporting data |
| --- | --- |
| **Theme:**<br>Describing personal experience | The overwhelming sentiment among teens with long Covid is a sense of loss. "*I have missed out on everything*," says [redacted HCP name]. (*The Guardian*, 10<sup>th</sup> August 2021)<sup>41</sup><br>"My teenager said '*we're in a half life, we didn't die and we haven't recovered*'." (*The Times*, 13<sup>th</sup> March 2021)<sup>30</sup><br>*'What is happening to me?'* The teenagers trying to make sense of long Covid (*The Guardian*, 10<sup>th</sup> August 2021)<sup>41</sup> |
| **Theme:** CYP thematic overlap with other actors<br>**Subtheme:**<br>Validation of Long Covid | While long Covid is a condition that generally affects older people, teens and children can - and do - become ill. (*The Guardian*, 10<sup>th</sup> August 2021)<sup>41</sup><br>Children who have contracted Covid-19 are reportedly suffering the effects of the virus for months afterwards, debunking widespread opinion that children wouldn't be hit as hard as older patients. (*The Mirror*, 1<sup>st</sup> November 2020)<sup>39</sup><br>Now a new study from King's College London reveals that long Covid isn't just a condition of adults but can also affect children and young people. (*The Daily Mirror*, 18<sup>th</sup> October 2021)<sup>44</sup> |
| **Theme:** CYP thematic overlap with other actors<br>**Subtheme:**<br>Invalidation of Long Covid | Researchers say the findings suggest long Covid is likely less of a concern among kids and adolescents than it is among adults (*Mail Online*, 1<sup>st</sup> October 2021)<sup>45</sup><br>Long Covid symptoms rarely persist beyond 12 weeks in children and adolescents unlike adults, new research suggests. (*PA Media*, 16<sup>th</sup> September 2021)<sup>27</sup> |
| **Theme:** Formerly very able | The previously sporty teenager - who played football and rugby for local teams - could not take more than a few steps without being overwhelmed with exhaustion. (*The Sun*, 7<sup>th</sup> February 2021)<sup>26</sup><br>Aberdeenshire teen [redacted CYP name] used to be an avid skier, competing across the country and overseas but, since catching Covid, she struggles to walk far or carry out simple tasks. (*The Daily Record*, 12<sup>th</sup> September 2021)<sup>25</sup> |
|  | Devastated parents have said their previously healthy kids are now confined to wheelchairs and too fatigued to attend school (*The Mirror*, 4<sup>th</sup> September 2021 <sup>46</sup>) |

All included articles were manually converted into plain text format using Notepad, and text not related to the body (such as suggestions for further reading) were removed if spotted at the beginning or the end of the text file. All media articles were loaded into the corpus tool ANTconc,^13^ which was utilised to facilitate the analyses.

### Data analysis

We used a modified social actor theory approach^14^ to Critical Discourse Analysis (CDA) in determining how media articles (re)produce knowledge of Long Covid in CYP within existing power structures. In CDA, discourse is viewed as an inherently social practice that is both reflective of and influential on public perception and power structures.^15,16^ Our approach to CDA maintains the fundamental purpose of producing systematic and reproducible problem-oriented investigation^16^ but focuses on a craftsmen perspective of methodology^17,18^ in integrating elements of thematic analysis. The critical angle taken is informed by the conceptual framework of epistemic injustice.^9^

Each article was examined using ANTconc^13^ to locate various actors: parents of CYP with Long Covid, HCPs, and CYP with Long Covid. HCPs were defined as medical clinicians as well as scientists and researchers addressed as doctors. Actors were identified in the text via KeyWord in Context search (KWIC) (for the list of search terms used and results yielded, refer to **SI 7 table**). As a result, the articles were not read in full. Focusing on the overall representation of actors as opposed to individual articles provided a broad overview and allowed the researcher to identify key information and common themes through cross-referencing. Each line was read and coded thematically and linguistically.

Themes were derived based on the Braun and Clarke’s steps for thematic analysis.^19^ The discursive elements of the research were conducted based on Baker’s corpus-driven approach to discourse analysis.^20^ The thematic diagram was iteratively constructed for each actor based on the developing understanding of identified themes. Themes selected for inclusion in the results were based on saliency, relevance to the research question, and alignment with the conceptual framework of epistemic injustice. When beneficial, the significance or uniqueness of findings were evaluated against the BE06, a reference corpus. The BE06 is a publicly available, one-million-word corpus of published written British English and is intended to be used as a representative sample.^21^ For a detailed explanation of each step undertaken in the analysis and the rationale behind each step, refer to **SI 8 table**.

### Patient and Public Involvement (PPI)

PPI helped to gain insight into stakeholder perspectives. The founder of a patient advocacy group for CYP with Long Covid which is now a leading charity is a PPI co-author on this paper (SM). She shared her experience in that role, as a person with Long Covid, and as a parent seeking care for her child. The research co-production process involved a review of potential research questions as well as an overview of the public contributor’s lived experience. The methodology and focus of the research were modified in light of this.

### Ethical considerations

We analysed publicly available media articles. Names referenced in quotes from the published media articles were redacted.

### Quality assurance

These findings were all initially single coded by the primary researcher by hand. Coding of lines was reviewed against the final codebook to ensure strict adherence to the definitions of codes. Single coding likely allowed for greater consistency in the coding process, as there was no potential for coding discrepancy. As a result, reliability is high. To enhance validity, the data for all codes and themes included in the results were reviewed by a second author (DC) to ensure that the data were well represented by these themes. DC provided input on the accuracy of the coding classifications and provided input on the quotes to highlight as examples. The coding manual and comprehensive thematic maps are included to allow the reader to determine validity and the degree of confidence to be placed in the findings **(SI 9 table and SI 10-12 figures)**. Initial themes were discussed with the other co-authors to enhance trustworthiness in findings.

## Results

Themes selected for inclusion in the results section below were based on saliency, relevance to the research question, and alignment with the theoretical framework.

### Parents

#### Parents versus mothers

Of the 181 times the actor parents were identified via the KWIC tool, 57 instances referred explicitly to the mother. No instances were found where the father of the CYP was specifically referenced. To determine if this is unique to the corpus created for this research, the search terms used were replicated in the BE06. The BE06 identified 1684 references to parents via the KWIC tool, of which instances referring explicitly to the mother and father were exactly equal. This indicates that the absence of fathers in this corpus is atypical. The presented themes collate mothers and parents, with specifications when themes between mothers and parents differed.

### Knowledge produced by parents

#### Parents providing information

The primary function of parents in the corpus was to recount symptoms or the experience of CYP with Long Covid. Often parents reported CYP’s symptoms in the context of contributing to research. Sometimes parent-reported symptoms were regarded neutrally, but they were frequently framed as a research limitation or a source lacking credibility. Parents’ reporting of CYP’s symptoms were devalued when they were seen as subjective and potentially exaggerated. Of note, symptoms are inherently subjective,^22^ so the criticism may more accurately reflect criticism of the diagnostic criteria which is based on symptoms and not biomedical markers.

#### Parents highlighting a lack of support and experiencing personal attacks

The media articles heavily featured parents highlighting a lack of support for themselves and CYP with Long Covid. Within the theme of a lack of support, parents reported minimisation of Long Covid in CYP. In addition, parents cited numerous barriers to care and a lack of available services. Parents also reported experiencing personal attacks in seeking support for their CYP with Long Covid. In many of these attacks, the credibility of the parent was questioned.

The media articles primarily presented lack of support and experiences of personal attacks through quotations and reporting of parents’ perception. Media articles did not present parents experience as factual. As a result, the responsibility for the accuracy of the claims lies within the referenced parents as opposed to the journalist. This created an opportunity to devalue parental accounts. For example, in writing “[redacted CYP name] and her mother [redacted parent name] feel the illness hasn’t been taken seriously”,^23^ using the word “feel” highlights their subjective perspectives as opposed to contextualising their experiences within evidence that Long Covid is indeed not taken seriously.^24^ In another example, “parents of children with the condition claim nothing has been done to help them”,^25^ the word “claim” alongside the extreme “nothing” implies that parents’ statements may be unreliable.

#### Parents describing gaps in collective understanding

Parents also described how gaps in collective understanding have impacted CYP with Long Covid. One mother described Long Covid in CYP as “Russian roulette” in reference to the unpredictability of who becomes afflicted.^26^ Parents reported feelings of invisibility for CYP with Long Covid, in part due to lack of recognition or proof of the disease.

### Knowledge produced about parents

#### Mother presented as having Long Covid

Mothers (but never parents) were sometimes presented as also having Long Covid. In many of these instances, mothers also described siblings who had Long Covid.

## HCPs

### Knowledge produced by HCPs

#### Rarity discourse

HCPs were often included in the articles discussing the prevalence of Long Covid in CYP. Of all instances identified in the corpus, 30% of the time HCPs quantified prevalence neutrally and 70% of the time HCPs subjectively appraised the rarity of Long Covid. CYP suffer fewer chronic conditions as adults,^33^ so the often-used comparison of prevalence across these groups is unlikely to provide a complete account of “rarity” relative to CYP. When attaching a value judgement, 19% of the time HCPs viewed Long Covid in CYP as not rare, and 81% of the time HCPs described Long Covid in CYP as rare. When calling the condition rare, HCPs frequently stated that this should be reassuring for concerned parents. Usually, the CYP with Long Covid were not addressed in this context, but sometimes it was recognised that rarity is not a consolation for those currently affected. One HCP stated "Long Covid seems to be rare in children, but it doesn’t matter. If it’s your child there needs to be a service for even one child with Long Covid.”^34^ This deviant example provides a subjective judgement on rarity while still recognising the impact of Long Covid on affected families.

#### Perceptions of disease validity

HCPs also offered their perceptions on the validity of Long Covid in CYP. In most occurrences that explicitly addressed disease validity, the HCP emphasised that the condition is important to take seriously. However, in some instances the manner in which validation was delivered could be interpreted as backhanded. In one remark, Long Covid is seen as a “side effect”^35^ as opposed to a distinct and legitimate condition.

In addition, HCPs engaged in pathologisation avoidance,^10^ where they hesitated to characterise the experiences of CYP as abnormal or requiring a diagnosis. Pathologisation avoidance was also located in the CYP lines, where one professor quoted in *The Daily Telegraph* noted “…just how common symptoms such as tiredness or headaches are in children and teenagers, regardless of whether they had Covid or not.”^36^

Pathologisation avoidance in the case of Long Covid in CYP may be a form of wrongful depathologisation as the diagnosis is important for receiving care.

Wrongful depathologisation could be observed in a *PA Media* article,

“Dr [redacted HCP name] of the MCRI and University of Fribourg said symptoms of long Covid were difficult to distinguish from those attributable to the indirect effects of the pandemic, such as school closures, not seeing friends or being unable to do sports and hobbies.”^27^

The implication that indirect effects of the pandemic could be erroneously conflated as Long Covid suggests that symptoms of Long Covid are normal aspects of life for CYP impacted by the pandemic.

In other instances, HCPs engaged in overpsychologisation (where they over-attributed Long Covid to mental illness) of Long Covid or they gave an alternate mental health diagnosis based the psychological symptoms of Long Covid. The media articles featured a mix of HCPs perpetuating versus challenging the overpsychologisation of Long Covid.

#### Difficulties with diagnosis

HCP’s also referenced difficulties with diagnosing Long Covid, especially with no confirmation of an initial covid-19 infection. Many media articles were published before a definition was created. Even when the case definition was created, HCPs faced difficulties, with a *Scotsman* article noting “Leading public health experts have warned it is underestimated, due to a lack of understanding of the post-viral condition among doctors. And there is no simple test.”^31^

### Knowledge produced about HCPs

#### HCPs as uninformed

Throughout the corpus, HCPs were characterised as uninformed. One paediatrician warned that “experts are still baffled by the long-term complications of the disease.”^23^ This lack of knowledge may come from both the novelty of the condition (a pragmatic, not inherently unjust barrier), and a societal lack of conceptual framework to understand Long Covid (an inherently unjust barrier)^9,11^. In other instances, HCPs were outwardly characterised as unjustly ignorant. A *Wales Online* article read, “Long COVID is a well-recognised condition in children but sadly, there’s still poor awareness among some medical professionals.”^37^ For either reason, HCP’s being uninformed appeared to contribute to negative experiences and created a formidable barrier to diagnosis.

## CYP

### Knowledge produced by CYP

#### Describing personal experience

The most significant aspect of the knowledge generated by CYP was its noticeable absence. While the discourse of the corpus revolved around this actor, CYP were mainly spoken for or about. In the few instances CYP directly produced knowledge, it mainly consisted of CYPs describing the personal impact of Long Covid and grieving the parts of their lives that have changed. CYP sometimes highlighted uncertainty of their condition and the difficulty making sense of what has happened to them.

### Knowledge produced about CYP

#### Overlap with other actor groups

Many of the lines identified for CYP were similar to the lines identified in the parents and HCPs actor groups. There were many lines highlighting a lack of support, mostly from the parent’s perspective but sometimes from HCPs and the writer of the media article. In addition, the validity of Long Covid was frequently discussed in the CYP lines. Unlike the HCP actor group, the statements of validity often came from the writer of the media article. In both the validation and invalidation of Long Covid in CYP, explicit references to adults were frequently employed. In statements that validated the condition, the emphasis was on explaining that Long Covid does not only affect adults. In statements that invalidated the condition, the severity of the CYP’s condition was regarded as not as serious as in adults.

#### Formerly very able

CYP were frequently described as formerly very able. The CYP was described by others, typically parents or the writer of the media article, as opposed to providing this information themselves.

Our findings culminate in table 5, where our main results are mapped onto a framework of epistemic injustice to demonstrate practical effects of the media discourse.

**Table 5.** Conceptual framework of findings.

| How each actor experiences or enacted testimonial injustice |  |  |
| --- | --- | --- |
| Parents | HCPs | CYP |
| Portrayed as gendered<br>Mothers presented as also having Long Covid<br>Parents accounts reported as unverified opinions<br>Parents report experiencing personal attacks in response to seeking care | Rarity discourse portrays CYP with Long Covid as outliers<br>HCPs combats testimonial injustice when validating Long Covid in CYP<br>HCPs invalidates Long Covid through pathologisation avoidance and overpsychologisation<br>Often seen as contributing to negative healthcare experiences | Lack of their accounts<br>Portrayed as formerly very able in an attempt to bolster authority |
| How all actors experience hermeneutical injustice |  |  |
| Parents and CYP describe feelings of invisibility as Long Covid in CYP is often unrecognised and unverified<br>HCPs seen as uniformed<br>Issues with diagnosis<br>Long Covid in CYP validated or invalidated in relation to adult Long Covid |  |  |

## Discussion

The aim of the study was to determine how UK media articles characterise LC in CYP. This was explored through identifying prominent actors via search terms. The thematic content and the discursive strategies employed in the articles were systematically identified and presented.

This research has demonstrated the ways in which media characterisations of Long Covid in CYP reflect and contribute to epistemic injustice. The media articles both report on instances of epistemic injustice and create them in the discursive strategies used by journalists. Some instances of epistemic injustice, such as when parents are wrongly accused of child abuse, are poignant. However, other examples of epistemic injustice, such as the use of a rarity discourse to reassure unaffected families, are nuanced. While each infliction of epistemic injustice may seem minor, the cumulative effect leads to pervasive marginalisation of affected individuals.

### Parents and testimonial injustice

Parents experienced testimonial injustice when they were featured as gendered, sick, and their accounts were reported as unverified opinions. Mothers are often responsible for care-seeking, and their familial contributions are reported on more frequently than for fathers.^47^ While it was unsurprising that mothers were disproportionately referenced, the absence of fathers on their own was striking. Featuring mothers and not fathers may reinforce gender stereotypes.^48^

Mothers experienced testimonial injustice in the manner they were presented as also having Long Covid. Due to deeply-rooted societal prejudice against ill people,^10^ presenting mothers as having Long Covid may create stigma.^8^ This prejudice may be compounded by a historical scepticism of defined as subjective more common in women, such as connective tissue disease, ME/CFS, and now Long Covid.^10,49,50^ The negative consequences of this are exacerbated when the unique knowledge held by mothers with lived experience of Long Covid is unrecognised, as was seen in the corpus. Presenting mothers as also having Long Covid raised concerns of bias or CYP mimicking mothers. These concerns were explicitly expressed, with a *Wales Online* article stating, “Parents’ perceptions of their own symptoms may have influenced their perception or reporting of their children’s symptoms.”^51^ This speculation has direct negative effects. In *PA Media,* a mother was featured who “was told by a doctor that her daughter was only ‘‘mimicking’’ her symptoms.”^32^ This resulted in denial of care for the CYP with Long Covid.

There are many reasons why Long Covid may appear in family clusters. Researchers have identified potential genetic correlations with Long Covid.^52–54^ Additionally, a family member’s diagnosis can increase awareness and lead to other family members being correctly diagnosed with Long Covid. However, media articles emphasised research that “indicate[s] the critical role of family context on prolonged symptoms following SARS-CoV-2 infection” which “highlight the need for caution in interpreting the causes of prolonged symptoms in SARS-CoV-2 infected individuals, especially children” (*Wales Online).*^51^ Of note, the referenced study^53^ acknowledges the potential genetic explanation, but this is misreported in the *Wales Online* media article^51^ and mentioned as an aside.

The articles also featured instances where parents were personally attacked when seeking care. While important to highlight these injustices, this may create anticipated stigma for other parents.^8^ Anticipated stigma was discussed during the co-production stage of this research, where the public contributor recounted multiple cases where parents felt unsafe seeking care for their CYP due to potential allegations of abuse. Both the PPI input and the media articles referenced parents being accused of having Munchausen by proxy, which is both a mental illness and a form of child abuse. The corpus featured an account of a mother having a multi-agency referral form against her, which implied that custody of the child was at stake. When parents read media articles detailing personal attacks with such grave consequences, they understandably may decline to “come forward” to seek care for their CYP. This perpetuates the invisibility of CYP with Long Covid and decreases the likelihood of the CYP receiving appropriate care.

### Healthcare professionals and testimonial injustice

HCPs quoted in the corpus often played a role in perpetuating testimonial injustice through rarity discourse, invalidation of Long Covid as a physical illness, and as the actor contributing to negative healthcare experiences in the form of dismissal, personal attacks, pathologisation avoidance, and overpsychologisation. HCP’s combatted testimonial injustice when validating Long Covid in CYP and highlighting the mistreatment of affected families.

While likely done to assuage fear, the rarity discourse from HCPs can perpetuate feelings of isolation for affected individuals. During the co-production of this research, it was discussed how the portrayal of Long Covid in CYP as rare contributes to feelings of confusion and self-blame for parents. In addition, the alleged rarity of Long Covid does nothing to help those already afflicted and may silence them through characterising them as outliers. The rarity discourse may lead to the underestimation of prevalence and the under-allocation of resources to address Long Covid in CYP.

Given historical privileging of the authority of HCPs, particularly doctors,^55^ their validation and invalidation of Long Covid in the media holds great weight. Journalists significantly influence the direction of this discourse through the selection of HCPs to interview and quote. In the corpus, specific HCPs with repeated and unequivocally expressed scepticism of Long Covid in CYP were frequently quoted.

HCPs invalidated Long Covid in CYP through pathologisation avoidance. Pathologisation avoidance has been used to destigmatize groups such as the neurodivergent community.^10^ However, pathologisation avoidance in the case of Long Covid in CYP may be a form of wrongful depathologisation as the diagnosis is important for receiving care. Wrongful depathologisation has been seen in both ME/CFS and obsessive compulsive disorder,^10^ and constitutes an epistemic injustice.^10,12,56^

In addition, there was an implicit narrative that Long Covid is “just fatigue”. One HCP stated that “most people will experience some level of post-viral fatigue at some point in their lives”,^39^ with another HCP noting "the good news is that the majority of young people who get chronic fatigue tend to get better with appropriate support.”^40^ By switching from the term Long Covid to describing fatigue, the articles framed Long Covid and fatigue as one in the same. As seen in the ME/CFS literature, fatigue from a chronic condition is often misconstrued as something everyone experiences and is subsequently trivialised.^10^

HCPs also engaged in testimonial injustice where they overpsychologised Long Covid or they gave an alternate mental health diagnosis. Long Covid has mental health elements that should be recognised and addressed, but the whole attribution of the illness to mental health cause harm.^57,58^ Long Covid is a predominantly a multi-system multi-symptom disease.^59^ Giving a psychological diagnosis as opposed to a Long Covid diagnosis can harm wellbeing, and may lead to neglecting the physical symptoms of Long Covid.^6^ In addition, a wrong diagnosis is a form of hermeneutical injustice where patients are less able to make sense of their experience.^12,60^ Misdiagnosing Long Covid as a mental illness hinders progress in understanding Long Covid and producing effective treatments.^57,61^

People with Long Covid may underreport mental health symptoms because they reasonably believe their testimony will be misunderstood.^8^ This belief may come from experience, as over 95% of people with Long Covid experiencing at least one form of stigma and over 75% report experiencing stigma often.^8^ Testimonial smothering and its negative consequences have also been recorded in ME/CFS and in domestic violence disclosures.^10,62^ It can result in poor patient experience and may harm progress in understanding the mental health aspects of Long Covid.^63^ In the corpus, HCPs both forwarded the overpsychologisation narrative and challenged it. In one example, an HCP challenged the narrative, stating that “Kids with long Covid are treated terribly. The failings of doctors on this is huge. Most still put it down to anxiety.”^31^ HCPs may be among the most effective voices in challenging the whole attribution of Long Covid to mental illness, given their professional expertise. However, the salience of individual HCP voices is greatly influenced by who the media chooses to approach and quote, and there may be a selective bias.

Lastly, HCPs perpetuate testimonial injustice through invalidating experiences of Long Covid. Trust in HCP’s ability to address Long Covid in CYP may be eroded in those experiencing and reading about invalidating healthcare interactions. This loss of trust has profound public health implications.^64^ Patient’s trust is an important indicator of care quality, and is associated with better outcomes, treatment adherence, and timely seeking of care, which are important for recoveries and cost-efficiency.^64^

### CYP with Long Covid and testimonial injustice

CYP may have experienced testimonial injustice in the lack of coverage of their voice and in the presentation of being formerly very able. Of note, it can be difficult to distinguish testimonial injustice in CYP from justified differential treatment based on an established understanding that CYP’s capacity and legitimate epistemic ability develop with age.^65^ However, being a CYP is often a heuristic for epistemic unreliability to a greater magnitude than appropriate.^65^

CYP were largely excluded from producing knowledge in the corpus and were instead spoken for or about. While many are too young or too sick to contribute to articles, it is likely that there are CYP with Long Covid interested sharing their knowledge. As seen in how knowledge on Long Covid was created on Twitter, people with Long Covid have expertise that needs to be viewed alongside the traditional, medical knowledge base.^2,22,66,67^ A potential alternate explanation is that media outlets did seek the opinions of CYP, but CYP declined to participate, potentially due to anticipated stigma (which the media contributed to).^8^

Journalists and parents attempted to counter invalidation and minimisation of Long Covid through presenting CYP as formerly very able. This mirrors the way patients with ME/CFS have been described in the media.^10^ Boer argued that this characterisation is proactively employed to prevent depathologisation.^10^ Being formerly healthy deflects the blame-the-victim trope and delineates a stark contrast between before and after the condition developed.^10^ In addition, the formerly very able characterisation may promote increased general interest, as the condition is seen as something that can happen to even the healthiest individuals.^10^

Characterising CYP as formerly very able highlights how significantly Long Covid affects lives, but it does not give the CYP agency. CYP’s previous ability is used to emphasise their current inability, and this may contribute to the continued speaking for CYP with Long Covid and the lack of coverage on knowledge produced by this group. In addition, the use of the formerly very able trope to bolster validity implies that Long Covid may be less valid in a CYP that was not formerly very able. This further stigmatises CYP with Long Covid that have a previous chronic illness or disability. Some chronic illnesses have been shown to be associated with an increased risk of Long Covid,^68^ and the Long Covid experiences of individuals with comorbidities are equally important to take seriously.

### Hermeneutical injustice across actors

All actor groups are harmed by the hermeneutical injustice seen in Long Covid in CYP. One mother emphasised the difficulty of having her child’s Long Covid unrecognised, saying "I think because [redacted CYP name] looks fine, people think she is OK but she’s really not.”^25^ At the broader level, another parent noted that there is a “whole wave of children who were never diagnosed but now have long Covid, who are just a bit invisible in the system.”^30^ A diagnosis, while sometimes stigmatising, provides a hermeneutical device for CYP to understand their experience.^10,12,69^ Without a clear way to make sense of their ongoing symptoms, one teenager explained that “we’re in a half life, we didn’t die and we haven’t recovered’.”^30^ This “middle ground” between recovery and death was one of the primary aspects of Long Covid identified on social media.^2,67,70^The idea that covid is “mild” if the individual is not hospitalised created a false dichotomy that ignores the reality of Long _Covid._2,67,70

Long Covid can only be diagnosed when there is a probable acute covid-19 infection. This presents a hurdle to a diagnosis and available care. One parent interviewed in *The Times* described how lack of testing hindered a Long Covid diagnosis, “we have heard so many times from doctors that it isn’t related to Covid. They wouldn’t do an antibody test, I felt that they wouldn’t even give it a try. You hear about all these long Covid clinics, but no kids can get in them.”^30^

One HCP outlined the broader public health implications of this, stating “My concern is what happens if we don’t get confirmation of infection. This is so important. It will have an impact and will increase inequalities.”^31^ If confirmation of infection is essentially required to access services, many CYP will be unfairly denied care.

A few HCPs in the corpus mentioned an additional barrier to diagnosis, with one HCP in the *Scottish Daily Mail* stating ’The problem with kids is that, unless it’s blindingly obvious, it’s difficult getting information out of them.”^35^ While this may be overstated for older CYP, this is a legitimate concern for younger CYP. Some symptoms of Long Covid, such as anxiety, may be difficult for a CYP to fully comprehend, let alone explain.^71^

CYP are inherently at a hermeneutical disadvantage within the adult-created healthcare system, as their unique understanding and experience of illness is projected onto an adult interpretive framework.^12^ Within the covid-19 pandemic, there was a systematic de-prioritization of children’s interests^72^. The media initially portrayed children as vectors of covid-19 instead of individuals at risk.^72^ With mounting evidence that children contract covid-19, the narrative morphed to how covid-19 in children is mild^72^. This narrative has been countered with evidence that children (with and without underlying conditions) can suffer severe acute covid-19. Now, the narrative that children do not get Long Covid is causing harm. Policy decisions related to the pandemic in general have failed to fully consider potential harms for CYP and the risks associated with infection (including the risk of Long Covid), and this has been described as a form of childism.^73^ This builds off a historic context where medical research and discourse focuses on adults who are seen as those primarily at risk of chronic conditions.^33^

Validating Long Covid through saying it is similar in CYP and adults fails to recognize the unique challenges of Long Covid in CYP. Invalidating Long Covid through claiming that Long Covid does not affect CYP as often or as severely as adults also constitutes hermeneutical injustice. Long Covid is not necessarily less severe in CYP than it is in adults. Regardless, the “hierarchy of suffering” is a problematic concept^74^ that downplays the unique challenges faced by CYP.

### Strengths and limitations

The corpus-based method of this study was both a strength and limitation. The use of concordance lines enabled the researcher to review all articles located in a comprehensive, systematic search of UK media articles. The findings are therefore likely representative of UK media focused on Long Covid in CYP. A limitation is that the researcher did not read each line in the context of the entire article. This may have resulted in contextual misunderstandings. The researcher sought to compensate for this through an extensive data familiarisation phase. As is standard in corpus research,^20^ when the context of an element in the sentence was unclear, a larger section of the file was read and included. This resulted in a variable amount of context surrounding each search term, which may have resulted in the overcoding of certain themes based on inclusion of additional sentences. The data was single coded, and the study’s validity would have been improved with a second coder’s input.

Additionally, a limitation of the search term method to identify actors is that actors were not located when they were referred to by personal pronouns or proper nouns. It is possible that the themes identified via search terms were systematically different from the themes around personal pronouns or proper nouns of the actors.

A notable strength of this research was the research being co-produced with people with lived experience of Long Covid and advocating for its recognition in children. This enabled the research questions to focus on what is impacting families of CYP with Long Covid. In addition, the use of the conceptual framework of epistemic injustice focused the research and facilitates comparison with related examples of epistemic injustice in healthcare.

## Conclusion

This study highlighted discursive practices employed by journalists that contribute to epistemic injustice. The study’s findings also indicate a pattern of HCPs dismissing and stigmatising families impacted by Long Covid in CYP. Future research should seek to understand how families with Long Covid feel about media characterisations, and how this impacts efforts to seek and receive care. While this study focuses on the experience of CYP with Long Covid, findings may be generalisable. Media reporting has been shown to contribute to epistemic injustice ME/CFS in a similar manner.^10^ Based on this study’s findings, the researchers have identified recommendations for future reporting of Long Covid in CYP. These recommendations may be relevant to improve practices in reporting on other diseases in CYP.

## Data Availability

All relevant data are within the paper and its Supporting information files.

## Acknowledgments

This research was modified from a dissertation completed through the University of Southampton’s MSc programme in Public Health.

## Supporting Information Captions

S1 Table. LexisNexis search strategy.

S2 Table. List of included and excluded publishers from Lexis Nexis.

S3 Figure. Political leaning of media articles.

S4 Figure. Style of media articles.

S5 Figure. Year media articles were published.

S6 Table. Characterisation of each publisher.

S7 Table. Search terms used for each actor.

S8 Table. Analyses process for study.

S9 Table. Codebook for actors.

S10 Figure. Map of themes for parents.

S11 Figure. Map of themes for HCP.

S12 Figure. Map of themes for CYP with Long Covid.

