## Supplementary figures and images for "A Critical Analysis of UK Media Characterisations of Long Covid in Children and Young People"

### SI 3

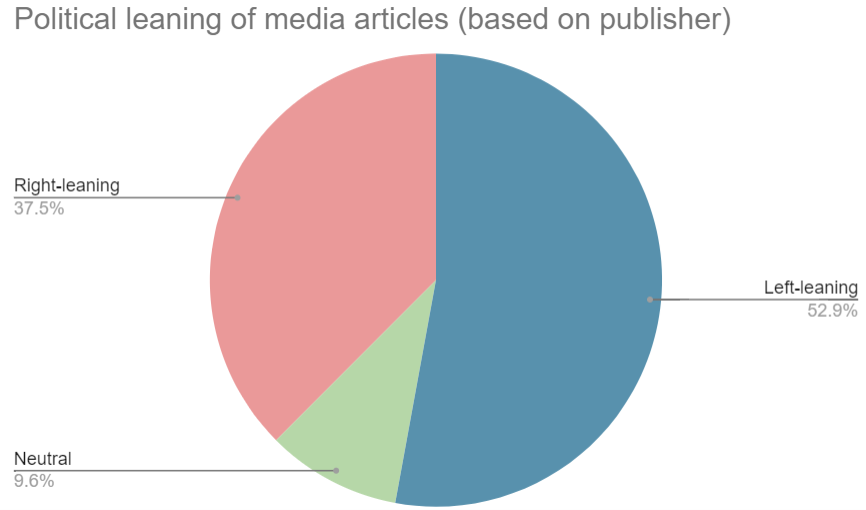

### SI 4

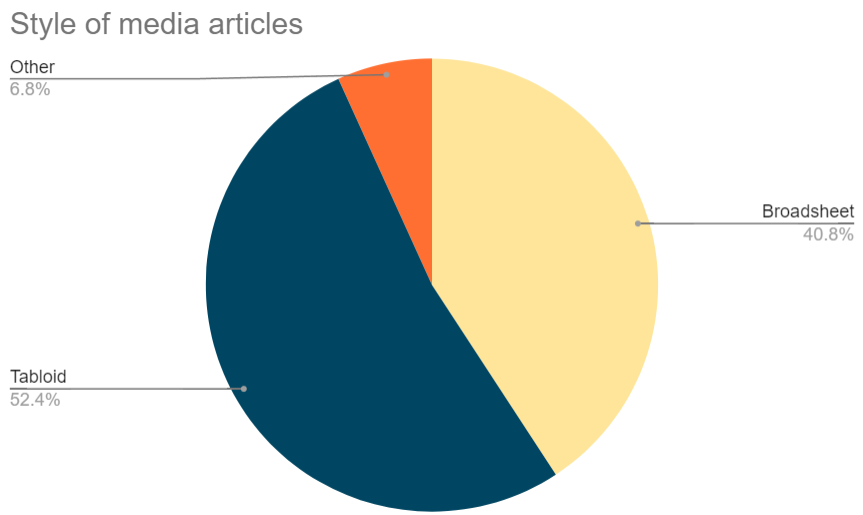

### SI 5

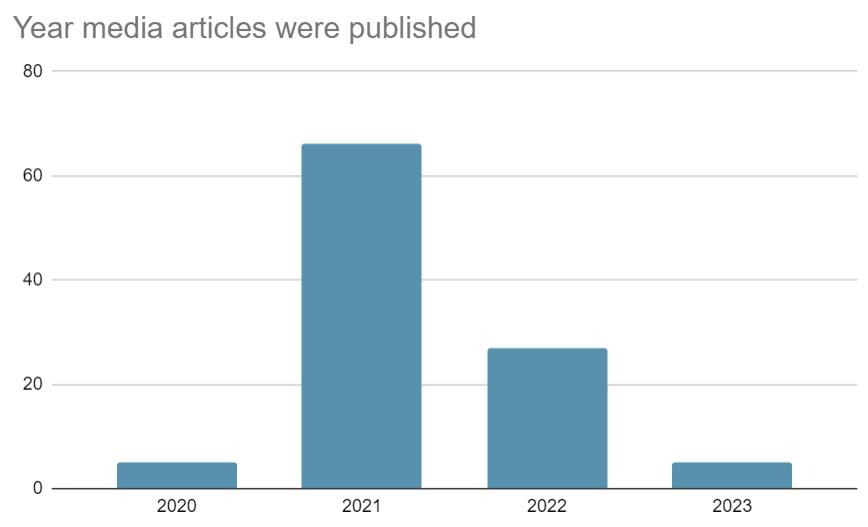

### SI 10

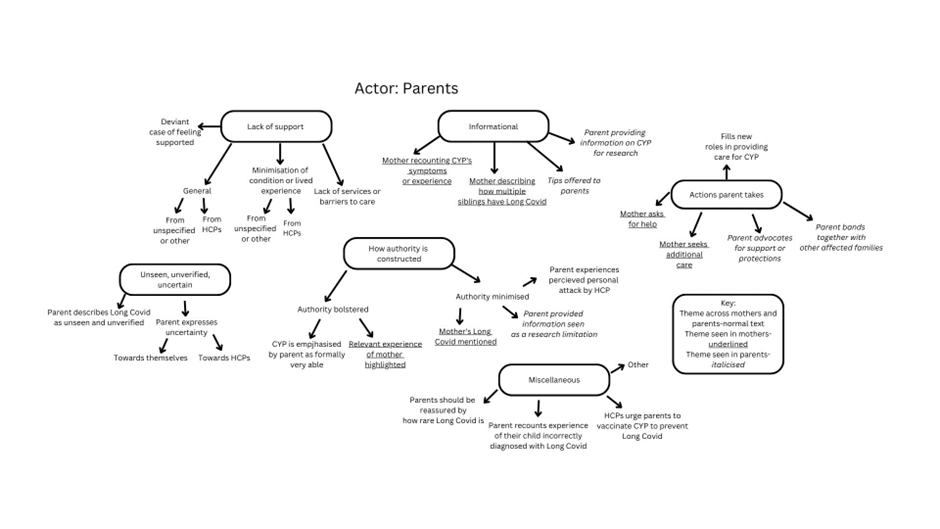

### SI 11

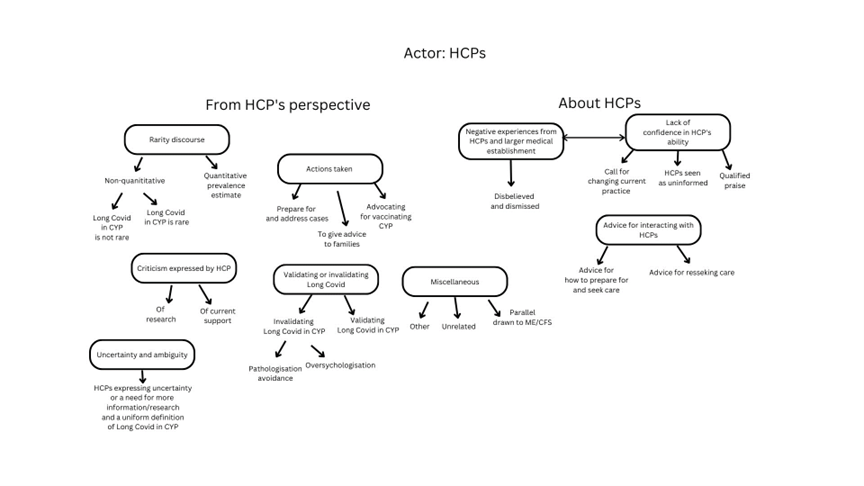

### SI 12

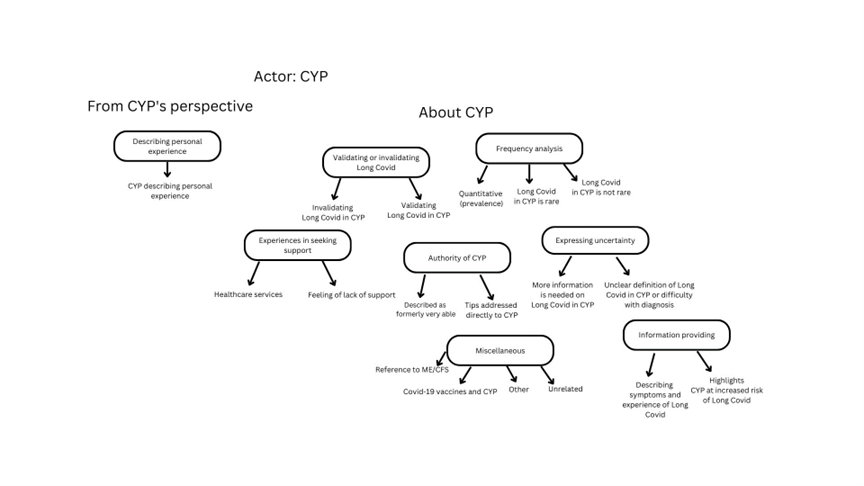
