## Supplementary material for "A Critical Analysis of UK Media Characterisations of Long Covid in Children and Young People": SI 6

**Supporting information 6. Characterisation of each publisher.**

| Name of publisher | Media style | Political leaning |
| --- | --- | --- |
| The Independent (United Kingdom) | Broadsheet | Left |
| MailOnline | Tabloid | Right |
| Daily Mirror/mirror.co.uk | Tabloid | Left |
| The Guardian (London) | Broadsheet | Left |
| The Daily Telegraph (London) | Broadsheet | Right |
| The Times (London)/thetimes.co.uk | Broadsheet | Right |
| PA UK and Ireland National Newswire | Newswire | Neutral |
| The Sun (England)/Thesun.co.uk | Tabloid | Right |
| i - Independent Print Ltd | Broadsheet | Left |
| Walesonline.co.uk | Tabloid | Left |
| The People | Tabloid | Left |
| Dailyrecord.co.uk | Tabloid | Left |
| The Daily Mail and Mail on Sunday (London) | Tabloid | Right |
| Scottish Daily Mail | Tabloid | Right |
| Herald Scotland | Broadsheet | Left |
| Daily Record and Sunday Mail | Tabloid | Left |
| The Express | Tabloid | Right |
| Scotsman | Broadsheet | Neutral |
| The Sunday Telegraph (London) | Broadsheet | Right |
| The Sunday Post | Tabloid | Neutral |
| Sunday Life | Tabloid | Right |
| The National (Scotland) | Broadsheet | Left |
| The Mail | Tabloid | Right |

*Publisher names as listed in LexisNexis. Source categories were drawn from various internet sources. Most publishers were indicated to have only a slight political bias. Note: the final list of included publishers is different from the earlier list of national publishers, as the list of national publishers was created prior to article screening.
