## Supplementary material for "A Critical Analysis of UK Media Characterisations of Long Covid in Children and Young People": SI 7

**SI 7. Search terms used for each actor.**

Boolean operators are recognised in ANTconc. The “*” symbol locates words that begin with the letters preceding the “*” operator. This allows the plural and singular form of words to be located. Searching a term in quotes directs the corpus tool to locate all instances of those words directly together in the same order.

| **Actor: Parents** | **Count** |
| --- | --- |
| Mother* | 23 |
| Mum* (includes “Mums and dads”) | 34 |
| Father* | 0 |
| Dad* (includes “Mums and dads”) | 2 |
| Parent* | 122 |
| “Mums and dads” | 2 |
| **Total** | **181** |

| **Actor: HCPs** | **Count** |
| --- | --- |
| Clinician* | 7 |
| Doctor* | 66 |
| Nurse* | 18 |
| “Healthcare professional*” | 5 |
| GP* | 41 |
| General practitioner* | 1 |
| Dr | 108 |
| **Total** | **246** |

| **Actor: CYP** | **Total Count in corpus** | **Count**  **analysed** |
| --- | --- | --- |
| Child* | 1000 | 50 |
| Young* | 218 | 50 |
| Kid* | 170 | 50 |
| Patient* | 86 | 50 |
| Teen* | 62 | 50 |
| Adolescent* | 36 | 36 |
| Infant* | 3 | 3 |
| Toddler* | 3 | 3 |
| **Total** | **1,578** | **292** |
