## Supplementary material for "A Critical Analysis of UK Media Characterisations of Long Covid in Children and Young People": SI 8

**SI 8 table.** **Analyses process for study.**

| Step | Action | Rationale |
| --- | --- | --- |
| 1 | Three key actor groups (HCPs, parents of CYP with Long Covid, CYP with Long Covid) were identified deductively based on background literature and PPI feedback. Several media articles were read in full to verify these are prominent actors in media portrayal of Long Covid in CYP. | Only three prominent actors were included in order to comprehensively examine each actor characterisation as opposed to conducting a superficial analysis of more actors. |
| 2 | Search terms for each actor group were identified | Search terms presented the most pragmatic means for identifying instances of the actor being referenced in the text. The use of search terms to identify discourses around actors was featured in Baker’s *Using Corpora in Discourse Analysis.*(Baker, 2023) |
| 3 | Through the KeyWord in Context (KWIC) function in ANTconc, concordances were generated each time a search term appeared in the text. Each parent and HCP concordance was read by the researcher for data familiarisation. For the CYP category, all search term with more than 50 occurrences had 50 concordance lines randomly selected for review (via the randomisation feature in KWIC). | Concordances list where a word occurs in a corpus alongside 10 words on either side of the search term. Concordance lines enabled the researcher to read the search terms in the context of the texts.(Baker, 2023) This stage constituted stage one of Braun and Clarke’s thematic analysis, data familiarisation.(Braun & Clarke, 2006) CYP concordance lines were not read in full as there were thousands more CYP concordance lines than the other actor groups. Randomly selecting 50 occurrences for each CYP search term resulted in more concordance lines than reviewed for the other two actors. This method for sampling concordances has been recommended by other researchers.(Baker, 2023) The unread concordance lines were randomly ordered and placed in a separate spreadsheet, where they could be reviewed if data saturation was not reached with the initial set of search terms and randomly selected lines. HCP and parents actor lines were read in full. |
| 4 | Each concordance was examined in the larger context of the article. The sentence (not the concordance) was extracted for analysis. If the sentence required the context of the previous or subsequent sentence to accurately preserve its meaning, those sentences were included in the abstraction. The data abstracts will now be referred to as lines. | For each actor, the lines were extracted for each search term in order to have all lines grouped into one of the three actor categories. Sentences were extracted as opposed to concordance lines because sentences typically included the actor and action while concordances often included the action of a previous sentence and the actor of a subsequent sentence without clear context. Additional sentences were included when they were necessary to accurately represent the context the search term was used in. |
| 5 | From reviewing the sentences and concordances, additional search terms to locate actors were identified. For the additional search term, steps 3-4 were repeated. | This was to ensure the search term lists were comprehensive. |
| 6 | Lines that included more than one search term within each actor category were located and marked to only be coded once (same line same article). Lines that were identical but in different articles or different parts of the same article were marked to be coded twice. | Identical lines repeated in the corpus were coded multiple times because the researcher sought to accurately represent salience. The same line identified through multiple search term searches was only included once as it only appeared once in the corpus. |
| 7 | Each line with a search term was reviewed, and initial thoughts were recorded. These initial thoughts referenced the content the lines as well as the discursive strategies and language used. | This was an additional stage of data familiarisation for the thematic and discourse analyses.(Baker, 2023; Braun & Clarke, 2006) |
| 8 | The initial thoughts and deductive insights were combined to create initial codes that were identified around each actor. These initial codes sought to capture (1) the knowledge shared by the actor and (2) how the actor is referenced by others. | Initial potential codes were identified deductively from searching published literature on Long Covid and on the often-compared condition ME/CFS, and from the PPI session. Literature on ME/CFS was included due to the parallels often drawn between the two conditions and the large body of discourse research on the condition. Initial codes were also identified inductively based on commonalities in language use and content across lines. This constituted stage two of thematic analysis, the initial generation of codes.(Braun & Clarke, 2006) |
| 9 | Each line was coded based on initial codes, with subsequent codes being added as additional elements were identified. Previous lines were reviewed when additional codes were added. Many lines were assigned multiple codes, but each element of the line was only coded for once. When lines were unclear, the researcher went back into the larger context of the article for clarity. | This furthered stage two of thematic analysis and was a later stage of generating codes for thematic and discourse analysis.(Braun & Clarke, 2006) The researcher did not restrict one code to one line because lines often included complex sentences with elements that could be coded separately. Each element was only coded once, as double coding the same element could lead to redundant themes and a misrepresentation of the saliency of certain codes or themes. |
| 10 | The list of initial codes was reviewed with some codes split or merged as the researcher refined codes and grouped them into broad themes. Themes were analysed to interpret how the language used shapes the themes and how the language is influenced by power dynamics in knowledge construction. | This was stage three of thematic analysis and an important step for discourse analysis,(Braun & Clarke, 2006) where the researcher combined codes into themes. Themes were developed both deductively from searching published literature on Long Covid and on ME/CFS, and from the PPI session. Themes were also developed inductively based on the codes generated in the data. |
| 11 | A thematic diagram was iteratively constructed for each actor based on the developing understanding of emerging themes. When necessary, previously coded data was re-examined in light of the newly articulated themes. | This was stage four of thematic analysis, the stage of reviewing themes where the relationship between themes and subthemes were considered in the context of each actor group.(Braun & Clarke, 2006) |
| 12 | The thematic diagrams were refined based on an understanding of the themes across the three actor groups. | This was stage five of thematic analysis, the defining of themes and subthemes in relation to the entire dataset.(Braun & Clarke, 2006) |
| 13 | The refined codes were reviewed to ensure accuracy. In addition, the refined codes were further analysed to identify discursive practices within themes. The broader sociocultural context of the data was considered in relation to the language used in the media articles. While reviewing the codes, the researcher also located key examples to represent themes and subthemes in the dataset. | This was a precursor to stage six of thematic analysis, the writing up of results.(Braun & Clarke, 2006) This also constituted a later stage of discourse analysis. |
| 14 | Key themes and subthemes were included in the results. Themes selected for inclusion in the results of this study were based on saliency, relevance to the research question, and alignment with the theoretical framework of epistemic injustice. | This constituted stage six of thematic analysis and an end stage of discourse analysis, locating examples and writing up the results.(Braun & Clarke, 2006) |
