## Supplementary material for "A Critical Analysis of UK Media Characterisations of Long Covid in Children and Young People": SI 9

### SI Table 9. Codebooks for actors.

**Parents**

| **Code** | **Subcode** | **Description** | **Example** |
| --- | --- | --- | --- |
| Lack of support | General, from HCPs | Parent feels a lack of support from HCPs | For the family it has been heartbreaking. For me as a mum to see her in so much pain and not being able to do anything, and having to fight the medical professionals |
| Lack of support | General, from unspecified or other | Parent feels a lack of support generally | 'It's terrifying': parents' struggle to get help for children with long Covid |
| Lack of support | Minimisation of condition or lived experience, from HCPs | Parent reports perception of HCP minimising condition or lived experience. This code includes cases of the parents reporting the HCP not taking the CYP or parents’ accounts seriously. This code also includes when parents report HCPs writing off Long Covid as a mental health condition | However, some parents say the condition is not being taken seriously by some GPs as most kids who catch Covid suffer no symptoms or long-term effects |
| Lack of support | Minimisation of condition or lived experience, from unspecified or other | This code encompasses when the parent expresses that Long Covid in CYP is not taken seriously enough. In this code, no HCP is explicitly mentioned | Despite the severity of her symptoms, Liliana and her mother Gail feel the illness hasn't been taken seriously |
| Lack of support | Lack of services or barriers to care | This code encompasses lack of services or barriers to care. This may include lack of vaccine access to prevent Long Covid, lack of services available, barriers to existing services | Parents tell the group they face a lack of support at every turn, from healthcare to support or children falling behind with school work |
| Lack of support | Deviant | This code encompasses all the deviant cases where parents express feelings of support from HCPs or in general. | Her mother, [redacted parent name], said clinicians have been supportive, but they have "openly admitted they don't know a lot about long Covid". |
| Informational | Parent recounting symptoms or the experience of their CYP with Long Covid | Parent providing anecdote of their CYP in media article | He also experienced rashes, bloodshot eyes and gastric pains, his mum said. |
| Informational | Parent providing information to researchers on Long Covid in CYP | Parent described as contributing to research, including in submitting symptom surveys | The Israeli survey, which was conducted between May and June 2021, found that 11.2 percent of parents said their children who recovered from the virus continued to experience symptoms. |
| Informational | Parent describes multiple children in house affected by Long Covid | Parent mentions that siblings are affected by Long Covid | A mum has 'changed forever' after watching her five children suffer a six-month 'long Covid' ordeal. |
| Informational | Tips are offered to parents | Media article offers tips to parents who have or could potentially have CYP with Long Covid | The course of the illness is not linear. While every child will have specific problems and individual needs, there are some suggestions we can make to help parents or guardians support their child or young person with long COVID |
| Unseen, unverified, uncertain | Uncertainty from parents about current situation or what to do | Includes parent expressing uncertainty about why their CYP had Long Covid, expressing uncertainty with what actions to take. Includes parents expressing uncertainty and worry for the future, and worry for potential future relapse | Her mother said: "For the family it has been heartbreaking. For me as a mum to see her in so much pain and not being able to do anything, and having to fight the medical professionals, I don't know what to do. She can't sleep at night. |
| Unseen, unverified, uncertain | Uncertainty towards HCPs | Parents feel that doctors do not have certainty on what to do or why Long Covid effects CYP | Her mother, [redacted parent name], said clinicians have been supportive, but they have "openly admitted they don't know a lot about long Covid". |
| Unseen, unverified, uncertain | Parent describes Long Covid as unseen and unverified | This includes cases where parents specifically reference feeling invisible or that Long Covid is unseen. This also encompasses cases where parents experience challenges with having Long Covid “verified” or formally diagnosed | The government insistence that children did not need to be tested means there is a "whole wave of children who were never diagnosed but now have long Covid, who are just a bit invisible in the system", said one parent. |
| Actions parent takes | Parent fills new roles in providing care for CYP | This includes new ways parent nurtures their CYP, how they take action to manage their CYP’s Long Covid, how they may give up work to better support their CYP | “..I am being her doctor, her nurse, her carer and her best friend. I just want to be her mum again." |
| Actions parent takes | Parent advocates for additional support or protections for CYP with Long Covid | This includes when parents demand new protections, the continuation of existing protections, access to vaccines, access to specialised clinics, etc | Parents are urging the government to set up specialist clinics to treat the "forgotten children" whose lives have been stalled by the virus. |
| Actions parent takes | Parent bands together with other affected families | This includes parents joining Long Covid patient advocacy groups | "It was only when I began doing some online research and found the Long Covid Kids support group that I realised there were other parents going through the same experience." |
| Action parent takes | Parent seeks additional care, reseeking same care or alternative or private care | When the parent reseeks the same type of care or private or alternate care after being unsatisfied with previous experience of seeking care. | Mum [redacted parent name], from Tallaght in Dublin, told The Irish Sun: "Because there are no consultancy plans in Ireland, we had to go to London two weeks ago and our trip cost us (EURO)3,000. |
| Action parent takes | Parent asks for help | Includes cases when parents appeal for help generally | The heartbroken mum of one of Scotland's youngest Long Covid patients has begged for help. |
| Constructing authority of parent | Authority minimised: Parent experienced perceived personal attack by HCP | When the parent recounts an instance when the HCP launched what could be considered a personal attack on the parent. This would include implying or saying the parent is neurotic, or implying a parent is mistreating their child | “…doctors have been dismissive to the point of telling me I'm an anxious mother and needed to calm down because children of my daughter's age are not affected by Covid or long Covid." |
| Constructing authority of parent | Authority minimised: Parent provided information seen as a research limitation | This includes cases when it is implied that the parents contribution is a limitation. This can be stated outwardly or more subtly referenced alongside other potential limitations. | He added: ``Current studies lack a clear case definition and age-related data, have variable follow-up times, and rely on self- or parent-reported symptoms without lab confirmation. |
| Constructing authority of parent | Authority minimised: Parent’s Long Covid mentioned | This encompasses cases where the parents own Long Covid diagnosis is included in the article | ``I look at all my children and none of them are the same children,'' she said. [Redacted parent name], a mother-of-two who has also been experiencing symptoms for seven months, added: ``We have no answers to this.'' |
| Constructing authority of parent | Authority bolstered: Relevant experience of the parent highlighted | This encompasses cases where a relevant aspect of a parent's experience is mentioned. This could be when the medical background of a parent is mentioned alongside the parents account of their CYP’s Long Covid. This could also include cases where the advocacy work of a parent is highlighted | Mum [redacted parent name], 37, a paediatric nurse, wants specialist help for him and the 3000 other children affected. |
| Constructing authority of parent | Authority bolstered: Parent emphasises CYP as formerly very able | This includes cases where the parent describes their CYP as formerly very able. This includes cases where the parent highlights the former academic and athletic achievements of their CYP | A mother from Lanarkshire said her 16-year-old son had been ‘left to rot’. [Redacted parent name] said: “ [Redacted CYP name] was a straight A student and now he sleeps 20 hours a day |
| Miscellaneous | Parents reassured by how rare Long Covid is | This encompasses cases that include an actor explicitly mentioning that the low prevalence of Long Covid should assuage the worries of parents | Long Covid is rare in children, according to results of a study that researchers hope will reassure parents and teachers. |
| Miscellaneous | Parent recounts experience of their child incorrectly diagnosed with Long Covid | This includes cases where a parent recounts an incorrect Long Covid diagnosis. All examples of this in the corpus refer to a case where a CYP with a brain tumour is misdiagnosed as having Long Covid | But despite being admitted to A&E, doctors mistakenly assumed he had migraines caused by long Covid, according to his mother. |
| Miscellaneous | HCP’s urge parents to vaccinate CYP to prevent Long Covid | This includes cases when an HCP discusses the importance of parents vaccinating CYP in order to prevent Long Covid | DOCTORS have appealed to parents to get their kids vaccinated against Covid after a poll showed just 25% planned to take up the offer of jabs for five to 11-year-olds next month. |
| Miscellaneous | Other | This includes any other instances where parents of CYP with Long Covid are referenced | One reason put forward as to why Covid was found to stay in family groups was genetics, suggesting parents and their children were predisposed to carry the virus. |
| Unrelated | Unrelated | This includes cases where the parent mentioned is not a parent of a CYP with Long Covid | Researchers said that studies have shown the pandemic has affected child development through the impact of isolation, poverty, food insecurity, loss of parents and caregivers, loss of time in education and stress. |

*Some quotes were repeated to illustrate examples where different aspects of the quote can be coded separately

**HCPs**

| Code | Sub-code | Description | Example |
| --- | --- | --- | --- |
| Rarity discourse by HCP | Quantitative prevalence estimate | The HCP neutrally estimates prevalence of Long Covid in CYP | "The Office for National Statistics found that after 12 weeks, 7% of two- to 11-year-olds, 8% of 12- to 16-year-olds and 11.5% of 17- to 24-year-olds had symptoms," said Dr [Redacted HCP name], an associate professor in public Health at the University of Southampton. |
| Rarity discourse by HCP | Non-quantitative value judgement, Long Covid in CYP is rare | The HCP appraises Long Covid in CYP as rare. This may be done in an attempt to reassure parents or after numerically estimating prevalence. This also includes cases when HCP say Long Covid in CYP is rarer than people think | Dr [redacted HCP name], consultant paediatrician at the UK Health Security Agency and study chief investigator, said: 'It is reassuring that the vast majority of primary and secondary school aged children surveyed since March 2020 have not experienced long Covid symptoms. 'These new data should be reassuring for parents, clinicians and policy-makers. |
| Rarity discourse by HCP | Non-quantitative value judgement, Long Covid in CYP is not rare | This encompasses the discordant cases when Long Covid in CYP is explicitly referred to as not rare. | The findings come after a separate report, published earlier in the week, said that children were unlikely to suffer from long Covid. Dr [Redacted HCP name] a scientific adviser at the National Institute for Health Research (NIHR), who was not involved in either study, said the ONS estimate suggests the condition is "hardly rare". |
| Actions taken by HCP | Prepare for and address cases | This includes cases when HCP’s prepare for and address cases. This also includes when HCPs quantify the number of patients helped | Dr [redacted HCP name], director of the Pediatric Post-Covid Program at the Children's National hospital, said they are booked out until March. She explained that over half of the program's 60 patients had come in just the last three months, despite opening in May 2021. |
| Actions taken by HCP | To give advice to families | This includes cases when the HCP directly addresses families to provide advice | I'm a GP and these are my top tips for what to do if you think you might have long COVID |
| Actions taken by HCP | Advocating for vaccination of CYP | This includes any cases where HCPs advocate for vaccination of CYP against covid-19 | Epidemiologist Dr [redacted HCP name] says vaccinating over-12s could help protect them against long Covid |
| Criticism expressed by HCP | Criticism of current support available to CYP with Long Covid | This includes criticism of other HCPs, criticism of the medical establishment’s response to Long Covid, criticism of current support measures | Dr [Redacted HCP name] said: "Kids with long Covid are treated terribly. The failings of doctors on this is huge. Most still put it down to anxiety. The narrative that most children are fine and  unaffected has been unchallenged. This puts parents in an impossible position, battling to be believed by healthcare practitioners and schools who often try to force children back. That doesn’t work. |
| Criticism expressed by HCP | Criticism of current research | This includes criticism of the research methods of studies on Long Covid in CYP | "I don't know that that is a very accurate way of gathering data ... I don't think that's likely to be truly reflective of how many young people are really experiencing persistent symptoms," said Dr [Redacted HCP name], senior clinical lecturer in paediatric infectious diseases and immunology at Imperial College London. |
| Perceptions of disease validity | Invalidating Long Covid in CYP- pathologisation avoidance | This includes cases where HCPs imply that many aspects of Long Covid may be the result of other factors, or are expected experiences in CYP | Dr [Redacted HCP name] said long Covid exhibits the same pattern as other post-viral illnesses, which children are as susceptible to, as adults. Most people will experience some level of post-viral fatigue at some point in their lives. |
| Perceptions of disease validity | Invalidating Long Covid in CYP- overpsychologisation | This includes cases when Long Covid is simplified to just refer to the psychological elements, or cases where alternate diagnoses of mental health disorders are suggested as opposed to a Long Covid diagnosis | The study, which has been running since March 2020, involved 134 schools and inputs from the parents of 4,870 pupils. Dr [Redacted HCP name], of King's College London, said: "There was no significant difference in the numbers presenting with a 'probable mental disorder' between both groups, whether test positive or negative. |
| Perceptions of disease validity | Validating Long Covid in CYP | This includes cases where HCPs focus on the importance of taking Long Covid seriously. This also encompasses cases where HCPs validate the experiences of CYP and their families, and discuss their poor treatment. Validating Long Covid also includes cases when invalidation of Long Covid is challenged, for example when HCPs criticise  overpsychologisation. | GP and author Dr [Redacted HCP name] said that while the virus was mild in young children, they were getting long Covid, which was a "real concern". |
| Uncertainty and ambiguity | HCPs expressing uncertainty or a need for more  information/research  and a uniform definition  of Long Covid in CYP | This encompasses HCPs highlighting the need for more information and the difficulties of having no uniform diagnosis for Long Covid in CYP | Dr [Redacted HCP name], a GP and Glasgow Tory MSP, has raised concerns that long Covid in children is that it can be particularly difficult to diagnose. He said: 'The problem with kids is that, unless it's blindingly obvious, it's difficult getting information out of them. |
| Other actors express negative experiences from HCPs and the larger medical establishment | Other actors report being disbelieved and dismissed by HCPs | This includes reports from other actors where HCPs disbelieve or dismiss a CYP’s Long Covid. This includes cases where Long Covid is written off as only psychological. | [Redacted parent name] is certain that her two teenage daughters both have long Covid. "I'm a senior nurse and I still feel patronised and dismissed by the paediatricians," she said. "Nothing has happened unless I've threatened to complain. |
| Lack of confidence in ability of HCP | Actors specifically note how HCPS are uninformed | The negative experience described by the actor includes an appraisal of HCPs as uninformed and therefore unable to address the needs of CYP with Long Covid | "Our children aren't being recognised as Long Covid sufferers because doctors aren't joining the dots between a wide range of symptoms. " |
| Lack of confidence in ability of HCP | Other actors call for change to improve ability of HCPs and medical establishment to address needs | An actor calls for more training/support/changing current practice due to the current inability of HCPs to address the needs of CYP with Long Covid | “Kids with long Covid can’t advocate for themselves so it's been a struggle to be heard. Healthcare professionals and support services need to know how to spot long Covid and properly support kids. They also need trained on what not do to. |
| Lack of confidence | Other actors express qualified support for HCPs | Actors express praise for HCPs in a way that recognises their effort but highlights their inability to help the CYP with Long Covid | "Because I can't really do much about it, I have to just kind of go with the flow and see what comes up." Her mother, [redacted parent name], said clinicians have been supportive, but they have "openly admitted they don't know a lot about long Covid". |
| Other actors provide advice for interacting with HCPs | Advice for preparing for and seeking care initially | Advice about HCPs is given to those seeking care for Long Covid in CYP | For example, if you've been feeling breathless, wear a loose-fitting shirt as the doctor will probably want to listen to your chest. |
| Other actors provide advice for interacting with HCPs | Advice for reseeking care following a negative experience | Advice for those reseeking Long Covid care is given. In this code there needs to be an indication that a previous attempt was made to seek care, but it was insufficient. | Talk to his doctor again. Make it clear your son is experiencing continuing symptoms and depressive thoughts. |
| Miscellaneous | Reference made to ME/CFS | Any reference made by or towards HCPs that created a parallel to ME/CFS | Dr [redacted HCP name], medical advisor for the ME Association, said: “We are, not surprisingly, receiving regular reports from people with ME/CFS who have had Covid and have then had a significant and prolonged exacerbation or relapse of their ME/CFS symptoms, which are very similar to long Covid.” |
| Miscellaneous | Other | Any instance in the corpus that involves an HCP in the context of Long Covid in CYP that is not otherwise addressed by existing codes | [Redacted HCP name], a paediatrician at the Gemelli University Hospital in Rome, led the first attempt to quantify long Covid in children. |
| Miscellaneous | Unrelated | Any instance of HCPs being mentioned unrelated to Long Covid in CYP | Dr [redacted HCP name], a senior clinical lecturer at the University of Exeter's medical school, said the bubble system has been good at limiting outbreaks. |

*Some quotes were repeated to illustrate examples where different aspects of the quote can be coded separately

**CYP**

| Code | Sub-code | Description | Example |
| --- | --- | --- | --- |
| CYP’s describing personal experience | CYP’s describing personal experience | Any instance where a CYP is describing their own experience of Long Covid, including symptoms, illness course, sense of loss, and sensemaking of their experience | A teenager suffering with Long Covid says at one point 'she thought she'd die' as her symptoms worsened. [Redacted CYP name], 16, said her symptoms included chest cramps and nerve pain which left her skin hurting if touched, to the point it "felt like someone was stabbing me". |
| Perceptions of disease validity | Invalidating Long Covid in CYP | This includes cases where the severity of Long Covid is invalidated. This could take the form of an actor saying that CYP recover from Long Covid quickly, or in the form of an actor saying Long Covid is not a concern in CYP, or an indication that CYP are pretending to have Long Covid, or pathologization avoidance. This also includes cases where Long Covid in CYP is seen as not much of a concern as Long Covid in adults | Prof [redacted HCP name], who is also a professor of paediatric infectious disease at the University of Melbourne and head of infectious diseases at The Royal Children's Hospital, said it was reassuring there was little evidence symptoms persisted longer than 12 weeks, suggesting long Covid might be less of a concern in children and adolescents than in adults. |
| Perceptions of disease validity | Validating Long Covid in CYP | This includes cases where Long Covid is explicitly validated or described in a way that “debunks” myths such as only adults get Long Covid. | Children and young people can also suffer from long COVID following even a mild infection with the virus. |
| Rarity discourse | Quantitative (prevalence) | This includes cases where the prevalence of Long Covid in CYP is quantified | The overall data collected during the seven months between September and March from teenagers who tested positive and may have had three or more symptoms 15 weeks after Covid, could be at least 4000 (best case scenario) and possibly as high as 32000 (worst case.) |
| Rarity discourse | Long Covid in CYP is rare | This includes cases where Long Covid in CYP is discussed as rare, or as rarer than people think | Very few children and teenagers infected with COVID-19 have long-term symptoms, a new study suggests. |
| Rarity Discourse | Long Covid in CYP is not rare | This includes cases where Long Covid in CYP is explicitly discussed as not rare | The world’s largest research to date involving children, the Children & Young People with Long Covid (CLoCk) study, found a significant number remain affected even 15 weeks after their infection. |
| Experiences in seeking support | Healthcare services | This encompasses cases where services are discussed. It includes when barriers to care are mentioned, additional support is advocated for, new support is detailed, support given is quantified or explained | Long Covid Kids is helping a family in Fife whose 13-year-old has been housebound for two years. The teen has not been able to get a wheelchair or school help. |
| Experiences in seeking support | Feeling of lack of support | This encompasses more of the psychosocial elements of seeking support. This includes cases when CYP are described as feeling forgotten or dismissed | And she said: "Kids are suffering from 'long Covid', there seems to be a lot of denial of that from the medical community and the governments and it's not helpful, the healthcare system is going to struggle." |
| Authority of CYP | CYP described as formerly very able | This includes cases where CYP are described as formerly very able | The previously sporty teenager - who played football and rugby for local teams - could not take more than a few steps without being overwhelmed with exhaustion. |
| Authority of CYP | Tips addressed directly to CYP | This includes cases when CYP are addressed directly with tips to handle Long Covid | Notably, many young people with long COVID probably haven't experienced living with a long-term illness, and so may not be familiar with navigating the healthcare system, which can be tricky. I'm a GP and these are my top tips for what to do if you think you might have long COVID. |
| Expressing uncertainty | More information is needed on Long Covid in CYP | This includes cases where an actor expresses the need for more information or better research on Long Covid | "There's hardly any evidence or data from around the whole world about long Covid in children and young people, and that's why it's vital that we do this research now," [redacted HCP name] said. |
| Expressing uncertainty | Difficulty of diagnosis or unclear definition of Long Covid in CYP | This includes cases where an actor expresses challenges with diagnosing Long Covid in CYP or the lack of a clear definition | There is no case definition of long Covid in children and young people in the way there is in adults. |
| Information providing | Describing symptoms and experience of Long Covid in CYP | This includes cases where CYP’s symptoms or experience are described | In the reviewed studies, the five most common long Covid symptoms reported in children and adolescents were headache, fatigue, sleep disturbance, concentration difficulties and abdominal pain. |
| Information providing | Highlighting CYP groups at increased risk | This encompasses cases where a certain risk group is highlighted | Teenage girls are most at risk of long Covid, according to a new study that revealed the effects have been found to run in families. |
| Miscellaneous | Parallel drawn to ME/CFS | This encompasses any case where ME/CFS is mentioned | Retired paediatrician [redacted HCP name] believes getting a diagnosis of CFS/ME is the best hope for children and young people with long Covid. |
| Miscellaneous | Referencing covid-19 vaccines and CYP | This includes any cases where CYP and covid-19 vaccines are mentioned. The CYP mentioned do not all have Long Covid, but vaccination to prevent Long Covid is mentioned | NHS advert uses young people with long Covid to promote jabs |
| Miscellaneous | Other | This includes any cases where CYP with Long Covid are discussed that is not encompassed by other codes | The number of young people suffering from long Covid must be considered when deciding to keep schools and universities open, a cross party panel of MPs has said. |
| Miscellaneous | Unrelated | This includes any cases where they search terms used and the lines are unrelated to CYP with Long Covid | To date, no Australian children have died of Covid-19. There has been one adolescent death, in a Sydney teenager who also had meningitis. |

*Some quotes were repeated to illustrate examples where different aspects of the quote can be coded separately
