## Supplementary material for "A Critical Analysis of UK Media Characterisations of Long Covid in Children and Young People": SI 1

**Supporting information 1 table. LexisNexis search strategy.**

| Search date: June 7^th^, 2023 in LexisNexis | Total |
| --- | --- |
| Headline("Long Covid" OR “Long-Covid” OR “Long Haul Covid” OR “Long-haul Covid” OR “chronic COVID” OR “Post Acute COVID-19 Syndrome” OR “Post Acute Sequelae of SARS CoV 2 Infection” OR “post covid condition” OR “post-covid condition” OR “post covid syndrome” OR “post-covid syndrome” OR PACS) | **46,826** |
| Headline(Kid OR Kids OR child OR children OR teen OR teens OR teenager OR youth OR “young people” OR adolescents OR minors) | **17,319,601** |
| Headline("Long Covid" OR “Long-Covid” OR “Long Haul Covid” OR “Long-haul Covid” OR “chronic COVID” OR “Post Acute COVID-19 Syndrome” OR “Post Acute Sequelae of SARS CoV 2 Infection” OR “post covid condition” OR “post-covid condition” OR “post covid syndrome” OR “post-covid syndrome” OR PACS) AND Headline(Kid OR Kids OR child OR children OR teen OR teens OR teenager OR youth OR “young people” OR adolescents OR minors) | **1,327** |
| Headline("Long Covid" OR “Long-Covid” OR “Long Haul Covid” OR “Long-haul Covid” OR “chronic COVID” OR “Post Acute COVID-19 Syndrome” OR “Post Acute Sequelae of SARS CoV 2 Infection” OR “post covid condition” OR “post-covid condition” OR “post covid syndrome” OR “post-covid syndrome” OR PACS) AND Headline(Kid OR Kids OR child OR children OR teen OR teens OR teenager OR youth OR “young people” OR adolescents OR minors)  Dates: January 1^st^, 2020- June 7^th^, 2023 | **1,239** |
| Headline("Long Covid" OR “Long-Covid” OR “Long Haul Covid” OR “Long-haul Covid” OR “chronic COVID” OR “Post Acute COVID-19 Syndrome” OR “Post Acute Sequelae of SARS CoV 2 Infection” OR “post covid condition” OR “post-covid condition” OR “post covid syndrome” OR “post-covid syndrome” OR PACS) AND Headline(Kid OR Kids OR child OR children OR teen OR teens OR teenager OR youth OR “young people” OR adolescents OR minors)  Dates: January 1^st^ , 2020- June 7^th^, 2023  Media: newspapers or newswires/press releases or magazines or webnews only | **817** |
| Headline("Long Covid" OR “Long-Covid” OR “Long Haul Covid” OR “Long-haul Covid” OR “chronic COVID” OR “Post Acute COVID-19 Syndrome” OR “Post Acute Sequelae of SARS CoV 2 Infection” OR “post covid condition” OR “post-covid condition” OR “post covid syndrome” OR “post-covid syndrome” OR PACS) AND Headline(Kid OR Kids OR child OR children OR teen OR teens OR teenager OR youth OR “young people” OR adolescents OR minors)  Dates: January 1^st^, 2020- June 7^th^, 2023  Media: newspapers or newswires/press releases or magazines or webnews only  Publisher: Europe, UK only | **405** |
| Headline("Long Covid" OR “Long-Covid” OR “Long Haul Covid” OR “Long-haul Covid” OR “chronic COVID” OR “Post Acute COVID-19 Syndrome” OR “Post Acute Sequelae of SARS CoV 2 Infection” OR “post covid condition” OR “post-covid condition” OR “post covid syndrome” OR “post-covid syndrome” OR PACS) AND Headline(Kid OR Kids OR child OR children OR teen OR teens OR teenager OR youth OR “young people” OR adolescents OR minors)  Dates: January 1^st^, 2020- June 7^th^, 2023  Media: newspapers or newswires/press releases or magazines or webnews only  Publisher: Europe, UK only  Publishers: National publishers only | **207** |
