## Supplementary material for "A Critical Analysis of UK Media Characterisations of Long Covid in Children and Young People": SI 2

**Supporting information 2. List of included and excluded publishers from Lexis Nexis.**

| Included publishers (prior to article screening) | Excluded publishers (prior to article screening) |
| --- | --- |
| The Independent (United Kingdom) *(24 articles)*  MailOnline *(22 articles)*  Daily Mirror/mirror.co.uk *(29 articles)*  The Guardian (London) *(14 articles)*  The Daily Telegraph (London) *(10 articles)*  The Times (London)/thetimes.co.uk *(12 articles)*  PA UK and Ireland National Newswire *(10 articles)*  The Sun (England)/Thesun.co.uk *(15 articles)*  i - Independent Print Ltd *(7 articles)*  Walesonline.co.uk *(7 articles)*  The People *(6 articles)*  Dailyrecord.co.uk *(6 articles)*  The Daily Mail and Mail on Sunday (London) *(5 articles)*  Scottish Daily Mail *(5 articles)*  Herald Scotland *(5 articles)*  Daily Record and Sunday Mail *(4 articles)*  The Express *(4 articles)*  The Western Mail (*3 articles*)  Scotsman *(3 articles)*  Scottish Express *(2 articles)*  The Sunday Telegraph (London) *(2 articles)*  The Sunday Herald (Glasgow) *(2 articles)*  The Scottish Mail on Sunday *(2 articles)*  The Sunday Post *(2 articles)*  Sunday Life *(1 article)*  The Sunday Express *(1 article)*  The National (Scotland) (*1 article*)  The Mail *(1 article)*  The Gazette *(1 article)*  The Leader *(1 article)* | Dorset Echo *(7 articles)*  Manchester Evening News *(5 articles)*  Standard.co.uk *(7 articles)*  Aberdeen Evening Express *(4 articles)*  Chronicle.live.co.uk *(4 articles)*  Gazettelive.co.uk *(4 articles)*  Birminghammail.co.uk *(4 articles)*  Evening Express *(4 articles)*  The Press and Journal *(4 articles)*  South Wales Argus *(3 articles)*  Daily Gazette *(3 articles)*  Grimbsy Telegraph *(3 articles)*  Swindon Advertiser *(3 articles)*  Nottingham Post *(3 articles)*  South Wales Echo *(3 articles)*  Getreading.co.uk *(4 articles)*  Irishmirror.ie *(3 articles)*  York Press *(3 articles)*  Huddersfield Daily Examiner *(2 articles)*  The Falmouth Packet *(2 articles)*  Lincolnshire Echo *(2 articles)*  Stoke the Sentinel *(2 articles)*  Aberdeen Press and Journal *(2 articles)*  Bucks Free Press *(2 articles)*  Daily Post (North Wales) *(2 articles)*  Hull Daily Mail *(2 articles)*  Basildon Echo *(2 articles)*  Liverpool Echo *(2 articles)*  Lancashire Telegraph *(2 articles)*  Leicester Mercury *(2 articles)*  The Argus *(2 articles)*  The Bolton News *(2 articles)*  St. Helens Star *(2 articles)*  The Northern Echo *(2 articles)*  Paisley Daily Express *(2 articles)*  Macclesfield-express.co.uk *(4 articles)*  Liverpoolecho.co.uk *(2 articles)*  The Journal (Newcastle, UK) *(2 articles)*  CoventryTelegraph.net *(4 articles)*  Coventry Evening Telegraph *(2 articles)*  The Express and Star *(2 articles)*  Shropshire star *(2 articles)*  The Evening Gazette *(1 article)*  Edinburgh Evening News *(2 articles)*  Birmingham Evening Mail *(1 article)*  Northumberland Gazette *(1 article)*  Kentish Gazette *(1 article)*  Wirral Globe *(1 article)*  News Shopper *(1 article)*  News Post Leader *(1 article)*  The Plymouth Herald *(1 article)*  Carmathen Journal *(1 article)*  Kent and Sussex Courier *(1 article)*  Birmingham Post *(1 article)*  Bournemouth echo *(1 article)*  Telegraph and Argus *(1 article)*  Braintree and Witham Times *(1 article)*  Bridport and Lyme Regis News *(1 article)*  Derby Telegraph *(1 article)*  East Kent Mercury *(1 article)*  Evening Times (Glasgow) *(1 article)*  East Anglian Daily Times *(1 article)*  Gloucestershire Echo *(1 article)*  Falkirk Herald *(1 article)*  Hereford Times *(1 article)*  Maldon and Burnham Standard *(1 article)*  Milton Keynes Citizen *(1 article)* Loughborough Echo *(1 article)*  Lancashire Evening Post *(1 article)*  Leigh Journal *(1 article)*  Tivyside Advertiser *(1 article)*  Scarborough Evening News *(1 article)*  Northwich Guardian *(1 article)*  Oxford Mail *(1 article)*  Peterborough Today *(1 article)*  Worcester News *(1 article)*  Runcorn and Widnes world *(1 article)*  The Sunday Mercury *(1 article)*  Western Daily Press *(1 article)*  Western Telegraph *(1 article)*  Airdrie & Coatbridge Advertiser *(1 article)*  Wishaw Press *(1 article)*  Dumfries & Galloway Standard *(1 article)*  The Royston Crow *(1 article)*  examiner.co.uk *(1 article)*  Royal Borough Observer *(1 article)*  Ayr Advertiser *(1 article)*  Ardrossan & Saltcoats Herald *(1 article)*  Irvine Times *(1 article)*  Impartial Reporter *(1 article)*  Helensburgh Advertiser *(1 article)*  Cumnock Chronicle *(1 article)*  North Wales Pioneer *(1 article)*  Isle of Wight County Press *(1 article)*  Owestry & Border counties Advertizer *(1 article)*  Chester Standard *(1 article)*  Denbighshire Free Press *(1 article)*  North Wales Chronicle *(1 article)*  Whitchurch Herald *(1 article)*  Dover Mercury *(1 article)*  Cambridge Independent *(1 article)*  Future News-Media Planner *(1 article)*  ENP Newswire *(6 articles)*  M2 PressWIRE *(4 articles)*  New Scientist *(1 article)*  The Week *(1 article)*  Pharmacy business *(1 article)*  Pediatric research *(1 article)* |
| Total included prior to screening: 207 | Total excluded prior to screening: 198 |

*Publisher names as listed in LexisNexis. All publishers were categorised as national (included) or not national (excluded) based on internet search of each publisher and review of categories by a librarian at the University of Southampton Highfield Campus.
